# Knowledge Discovery with Electrocardiography Using Interpretable Deep Neural Networks

**DOI:** 10.1101/2022.11.01.22281722

**Authors:** Lei Lu, Tingting Zhu, Antônio H. Ribeiro, Lei Clifton, Erying Zhao, Antonio Luiz P. Ribeiro, Yuan-Ting Zhang, David A. Clifton

## Abstract

Despite the potentials of artificial intelligence (AI) in healthcare, very little work focuses on the extraction of clinical information or knowledge discovery from clinical measurements. Here we propose a novel deep learning model to extract characteristics in electrocardiogram (ECG) and explore its usage in knowledge discovery. Utilising a 12-lead ECG dataset (*n*_ECGs_ = 2,322,513) collected from unique subjects (*n*_Subjects_ = 1,558,772) in primary care, we performed three independent medical tasks with the proposed model: (*i*) cardiac abnormality diagnosis, (*ii*) gender identification, and (*iii*) hypertension screening. We achieved an area under the curve (AUC) score of 0.998 (95% confidence interval (CI), 0.995-0.999), 0.964 (95% CI, 0.963-0.965), and 0.839 (95% CI, 0.837-0.841) for each task, respectively; We provide interpretation of salient morphologies and further identified key ECG leads that achieve similar performance for the three tasks: (*i*) AVR and V1 leads (AUC=0.990 (95% CI, 0.982-0.995); (*ii*) V5 lead (AUC=0.900 (95% CI, 0.899-0.902)); and (*iii*) V1 lead (AUC=0.816 (95% CI, 0.814-0.818)). Using ECGs, our model not only has demonstrated cardiologist-level accuracy in heart diagnosis with interpretability, but also shows its potentials in facilitating clinical knowledge discovery for gender and hypertension detection which are not readily available.

## 1 Introduction

AI-driven approaches, and in particular deep learning, are developing at pace, have increasing potential for application in healthcare, and have been used to address challenges for a variety of medical conditions [1], such as circulatory failure prediction [2], pulmonary tuberculosis testing [3], retinal disease diagnosis [4], and mortality prediction [5, 6]. In particular, encouraging developments in deep neural networks (DNN) have shown dermatologist-level classification of skin cancer [7], radiologist-level accuracy in identifying breast cancer [8], and ophthalmologist-level performance in detecting diabetic retinopathy [9]. The use of AI in healthcare has the potential to deliver meaningful impacts, both in improving productivity and efficiency of clinical practice, optimise workflow of care delivery, and in reducing medical errors through comprehensive diagnosis [10].

Despite the great promise of AI techniques in healthcare, concerns over the unknown interpretation process, i.e., blackbox model, have spurred a movement toward building trust in machine learning (ML) algorithms [11]; In particular, there are growing calls for transparent and trustworthy AI models from clinicians, lawmakers, and government regulators [1, 12]. For example, the European Union’s General Data Protection Regulation states that all people have the right to know meaningful information about the logic behind automated decisions using their data [10]; the U.S. Food and Drug Administration (FDA) emphasises the importance of interpretability among a set of terms for AI/ML practice [13, 14]. Transparency can support a physician’s competence in interpretation, and build trust within the physician–patient relationship; Conversely, a lack of this interpretive ability may impede the general acceptance of AI techniques in healthcare practice [12]. In addition, the improvement in interpretability of clinical data allows physicians to better understand the biological mechanisms behind disease, to identify disease-specific features, and enables efforts with the potential to derive more reliable biomarkers [12, 15, 16].

There are several efforts to develop interpretable techniques to produce explanations for ML decisions; such methods include class activation mapping (CAM) [17], local interpretable model-agnostic explanation (LIME) [18], Shapley additive explanations (SHAP) [19], and gradient-weighted class activation mapping (Grad-CAM) [20]. In particular, Grad-CAM and its variants have shown promising interpretation ability in processing medical images. For example, it was used to localise salient areas in chest radiographs for acute respiratory distress syndrome (ARDS) diagnosis [21]; segment chest X-ray images for COVID-19 detection [15], and identify scaphoid fractures in radiographic images [22]. However, these studies either focus on specific tasks or have limited experimental validation, and the interpretation capabilities of these techniques are still largely unexplored.

In this work, we hypothesise that AI models with a specific design can provide interpretation of healthcare data, identify clinically useful information, and facilitate discovering new knowledge that can be understood by clinicians. To test this hypothesis, we created, trained, and validated a novel AI model with a large-scale dataset; and we particularly focused on the interpretation of electrocardiogram (ECG), which is primarily due to the following two reasons. On one hand, ECG recording is the most commonly performed diagnostic test to screen cardiovascular diseases (CVD), which are responsible for more than 30% of all deaths globally [23]. It is understood that ECG recording provides an assessment of overall rhythm and cardiovascular status [24]; nevertheless, interpretation of the test varies greatly, even among cardiology specialists. Such variance between physicians presents a challenge to ensure consistency and reliability in the diagnosis; Moreover, the physician’s recognition of abnormal morphologies is mostly limited to existing cardiac disorders, it is therefore difficult to detect rare or relatively unknown diseases or recognise visually imperceptible elements in the morphology. On the other hand, modern technologies are constantly increasing the ability to acquire large numbers of ECG recordings, such that more than 300 million ECGs are obtained annually worldwide [25]. The data-intensive nature of ECGs requires comprehensive analytical methods to perform automated interpretation, which will facilitate understanding of the complexity of underlying diseases and ultimately improve healthcare outcomes.

There are recent studies showing advances in using AI techniques for digital ECG analysis, such as abnormal heart rhythm detection [26], cardiac contractile dysfunction identification [27], aortic valve stenosis screening [28], and early diagnosis of low ejection fraction [29]. However, most of these AI models focus on task performance rather than extracting clinically useful information or expanding knowledge from ECG recordings. For example, the AI model used in the study [26] demonstrated cardiologist-level detection of cardiac arrhythmia using ECGs; but the model primarily outputs diagnostic scores instead of explaining how the ECG morphologies were used to diagnose arrhythmias. From a clinical perspective, ECG morphologies characterise cardiovascular status and are used to derive disease-specific features for the diagnosis of arrhythmias, e.g., abnormal P waves for the diagnosis of atrial fibrillation [30]. Despite the impressive performance of AI models, it is unreasonable for either a patient or medical professional to accept an automated diagnosis at face value without justification [31]. More importantly, AI techniques are often highly complex, and thus require a substantial number of samples to train the model, without which outputs may be unreliable and have potential pitfalls [32]. For example, a treatment recommendation with an explicit contraindication could be made even by well-trained AI systems [10], but without an accompanying means of alerting the treating physician of the potential risk, there may be a consequence of major harm to patients.

Using our proposed interpretable DNN model, we perform three independent medical tasks with state-of-the-art diagnosis performance in this study, and in particular, the model enables to produce lead-specific interpretation of standard 12-lead ECG recordings. We first use the developed DNN model to identify and interpret rhythm abnormalities; this is because arrhythmias confer a substantial risk of mortality and morbidity in patients with heart failure, which represents a major healthcare burden and affects an estimated 64.3 million people worldwide [33]. Other than the diagnosis of heart conditions, we test the developed DNN model in a more general task of gender identification. This is highly relevant to our central task, because gender differences have been observed in the development of CVD; for example, women tend to develop heart disease later in life than men, while also having worse outcomes and higher mortality [34, 35]. In a further step, we perform the third task of hypertension screening to validate the developed model in a wider context of medical practice. Hypertension is the largest single contributor to CVD, causing stroke, heart failure, and coronary artery disease [36]. In particular, it has a rising prevalence and affects approximate 1.38 billion people worldwide (31.1% of the global adult population) [37]. To the best of our knowledge, this is the first time that an explanatory DNN model has been deployed and extensively studied with an ECG dataset of such a sheer scale. In particular, we identify salient features to explain the decision-making in each of the three tasks, and these features are also used to extract clinically useful information in the ECG recordings, i.e., dominant leads. We then further investigate the effectiveness of the identified dominant leads by performing various comparison studies in the three medical tasks.

## 2 Results

### 2.1 Overview and Study Population

Our developed explanatory deep learning model has two major components: a new architecture with channel-wise deep residual networks (CResNet) to implement the medical diagnosis, and an interpretation module to produce salient features that have been used for the decision-making. To validate the diagnostic and interpretation abilities of the developed model, we perform three independent medical tasks using a large dataset consisting of standard 12-lead ECG recordings (*n*_ECGs_ = 2,322,513) collected from unique individuals during clinic visits (*n*_Subjects_ = 1,558,772). Figure 1 depicts the dataflow and study population for the three medical tasks in this study; (*i*) In the first task, we use 2,315,782 ECG recordings to train the CResNet model to diagnose morphological abnormalities, including the first-degree atrioventricular block (1dAVb), right bundle branch block (RBBB), left bundle branch block (LBBB), sinus bradycardia (SB), atrial fibrillation (AF), and sinus tachycardia (ST). The trained CResNet model is then tested on a holdout ECG dataset, which is rigorously annotated by certified cardiologists; (*ii*) In the second task, we train the CResNet model for gender identification using ECGs collected from 1,398,907 subjects (female: 59.78%, *n*_Female_ = 836,267), which is tested with holdout ECGs sampled from 155,435 subjects (female: 59.52%, *n*_Female_ = 92,513); (*iii*) In the third task, the model is trained to screen hypertension for 1,398,907 subjects (hypertension: 31.66%, *n*_Hypertension_ = 442,918), and tested with 155,435 subjects (hypertension: 31.65%, *n*_Hypertension_ = 49,202). In both the second and third tasks, we select only one ECG recording for each individual, and when a subject has multiple ECG recordings, the earliest record is used.

**Figure 1:**
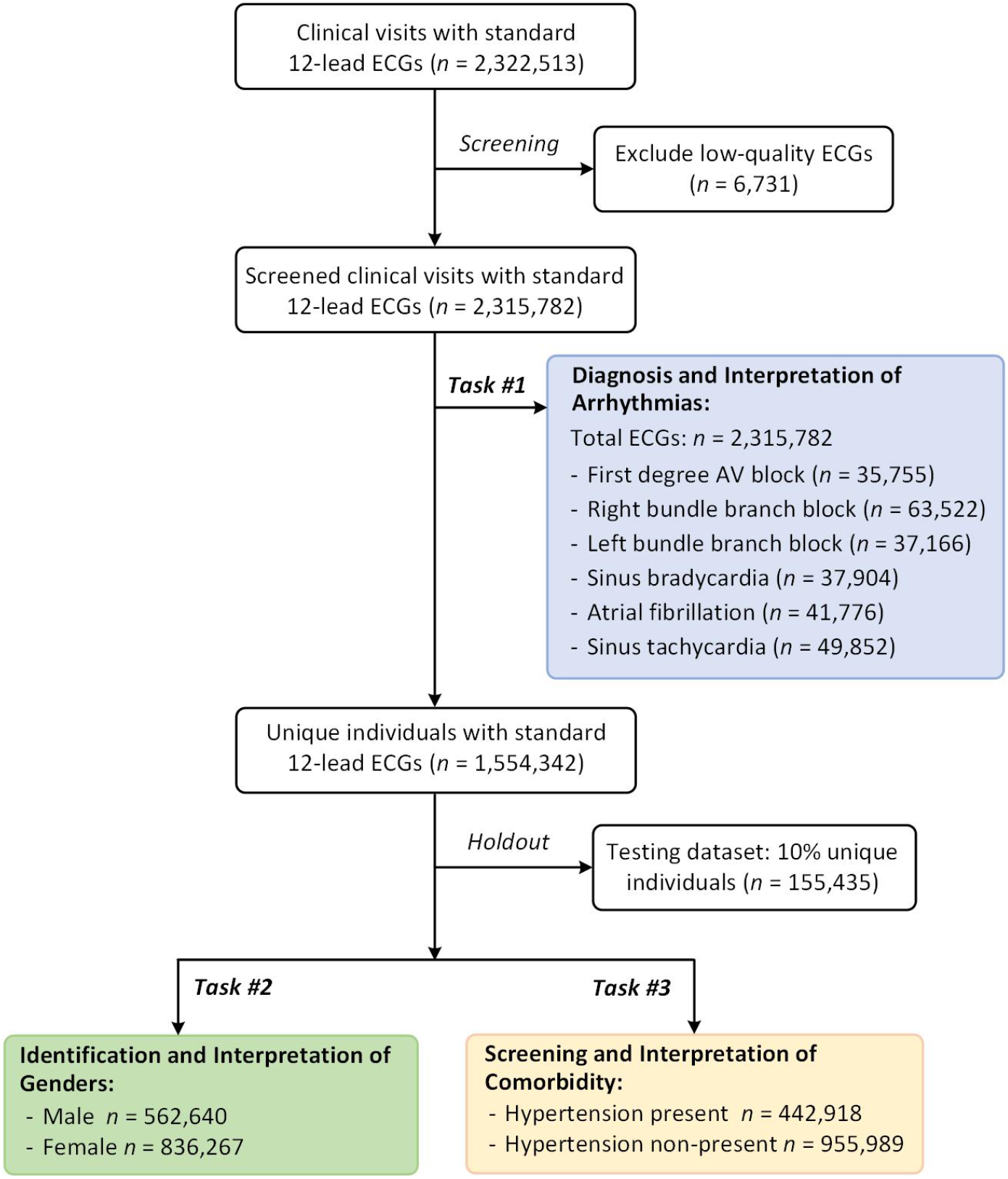
Dataflow and population characteristics for the three medical tasks in this study. The data was collected in primary care facilities with population characteristics as following. (*i*) For the first task (Task #1), the ECG recordings were collected from a population with an average age of 53.64 ± 17.42 years old (*yr*), and 60.26% are females (*n*_Female_ = 1,395,461); (*ii*) For the second task (Task #2), the population has an average age of 51.66 ± 17.58 *yr* and 59.78% are females (*n*_Female_ = 836,267); (*iii*) For the third task (Task #3), subjects with hypertension accounts for 31.66% of the whole population; the hypertension group has an average age of 59.33 ± 14.79 *yr*, and 62.71% are female subjects (*n*_Female_ = 277,756). Detailed descriptions of the dataset and population characteristics can be found in Extended Figure S1 and Table S1.

### 2.2 Diagnosis and Interpretation of ECG Morphological Abnormalities

In the first task, the CResNet model has a micro average AUC score of 0.998 (95% CI, 0.995-0.999) and an F1-score of 0.948 (95% CI, 0.921-0.971) on identifying the ECG morphological abnormalities. We report the F1-scores in Table 1A and compare the performance of our CResNet model with evaluation results from three junior professionals with experience in ECGs, two senior cardiologists, and the state-of-the-art study [38]. It can be seen from Table 1A that the highest evaluation score from the three junior professionals is 0.876 (95% CI, 0.830-0.915); The two senior cardiologists have higher performance than the junior professionals, with the highest F1-score of 0.945 (95% CI, 0.914-0.970); While the state-of-the-art benchmark [38] has a score of 0.938 (95% CI, 0.910-0.961). In comparison to the evaluation results yielded by the cardiology professionals, our CResNet model has better performance than the three junior professionals and one senior professional in the diagnosis of 1dAVb (*p* = 0.0433, in Extended Table S2). Furthermore, it significantly outperforms the three junior professionals in the diagnosis of AF (*p* = 0.0412), and has comparable performance with that of the senior cardiologists (*p* = 0.2482). We also provide the Cohen’s kappa coefficients to demonstrate the inter-rater agreement between our diagnosis with the evaluation results from the cardiology professionals in Extended Table S3. To show a comprehensive comparison of model performance, we present the evaluation results from our CResNet model, cardiology professionals, and the benchmark [38] in Figure 2. The highlighted symbol in the Figure 2 indicates the F1-score for each of the evaluation results, and the point at top-right corner of the figure is the ideal F1-score for the diagnosis. It can be seen from Figure 2 that the CResNet model has superior or comparable performance with evaluation results from the comparison studies, suggesting effectiveness of our model on the diagnosis of ECG abnormalities.

**Table 1A:** Performance comparison for the diagnosis of abnormalities using standard 12-lead ECG recordings.

|  | F1-score (95% CI) |  |  |  |  |  |  |
| --- | --- | --- | --- | --- | --- | --- | --- |
|  | Junior Professionals |  |  | Senior Professionals |  | DNN models |  |
|  | Cardio. Rd. | Emerg. Rd. | Medical Sd. | Cardio. #1 | Cardio. #2 | DNN_Comp [38] | CResNet |
| 1dAVb | 0.776<br>(0.625-0.889) | 0.719<br>(0.578-0.831) | 0.732<br>(0.605-0.836) | 0.828<br>(0.704-0.925) | 0.926<br>(0.844-1.000) | 0.897<br>(0.793-0.969) | 0.966<br>(0.912-1.000) |
| RBBB | 0.917<br>(0.842-0.974) | 0.852<br>(0.746-0.939) | 0.928<br>(0.852-0.985) | 0.957<br>(0.899-1.000) | 0.971<br>(0.921-1.000) | 0.944<br>(0.881-0.989) | 0.971<br>(0.928-1.000) |
| LBBB | 0.947<br>(0.875-1.000) | 0.912<br>(0.828-0.980) | 0.915<br>(0.830-0.983) | 0.966<br>(0.907-1.000) | 1.000<br>(1.000-1.000) | 1.000<br>(1.000-1.000) | 0.949<br>(0.882-1.000) |
| SB | 0.882<br>(0.743-0.976) | 0.848<br>(0.692-0.963) | 0.750<br>(0.538-0.889) | 0.897<br>(0.750-1.000) | 0.897<br>(0.741-1.000) | 0.882<br>(0.750-0.976) | 0.865<br>(0.727-0.973) |
| AF | 0.769<br>(0.545-0.933) | 0.696<br>(0.400-0.875) | 0.706<br>(0.500-0.865) | 0.870<br>(0.667-1.000) | 0.889<br>(0.737-1.000) | 0.870<br>(0.667-1.000) | 1.000<br>(1.000-1.000) |
| ST | 0.882<br>(0.789-0.951) | 0.946<br>(0.881-0.989) | 0.873<br>(0.778-0.949) | 0.896<br>(0.813-0.965) | 0.930<br>(0.853-0.987) | 0.960<br>(0.904-1.000) | 0.933<br>(0.862-0.987) |
| Micro_avg | **0.876**<br>**(0.830-0.915)** | 0.846<br>(0.793-0.892) | 0.833<br>(0.789-0.876) | 0.908<br>(0.871-0.941) | **0.945**<br>**(0.914-0.970)** | 0.938<br>(0.910-0.961) | **0.948**<br>**(0.921-0.971)** |

\*Cardio. Rd. — 4<sup>th</sup> year cardiology residents; Emerg. Rd. — 3<sup>rd</sup> year emergency residents; Medical Sd. — 5<sup>th</sup> year medical students; Cardio. #1 — the 1<sup>st</sup> cardiologist; Cardio. #2 — the 2<sup>nd</sup> cardiologist; DNN\_Comp — the state-of-the-art benchmark model developed in [38]; CResNet — the DNN model developed in this study. The bold-faced scores denote the best performance for the three junior professionals, two senior professionals, and two DNN models.

**Figure 2:**
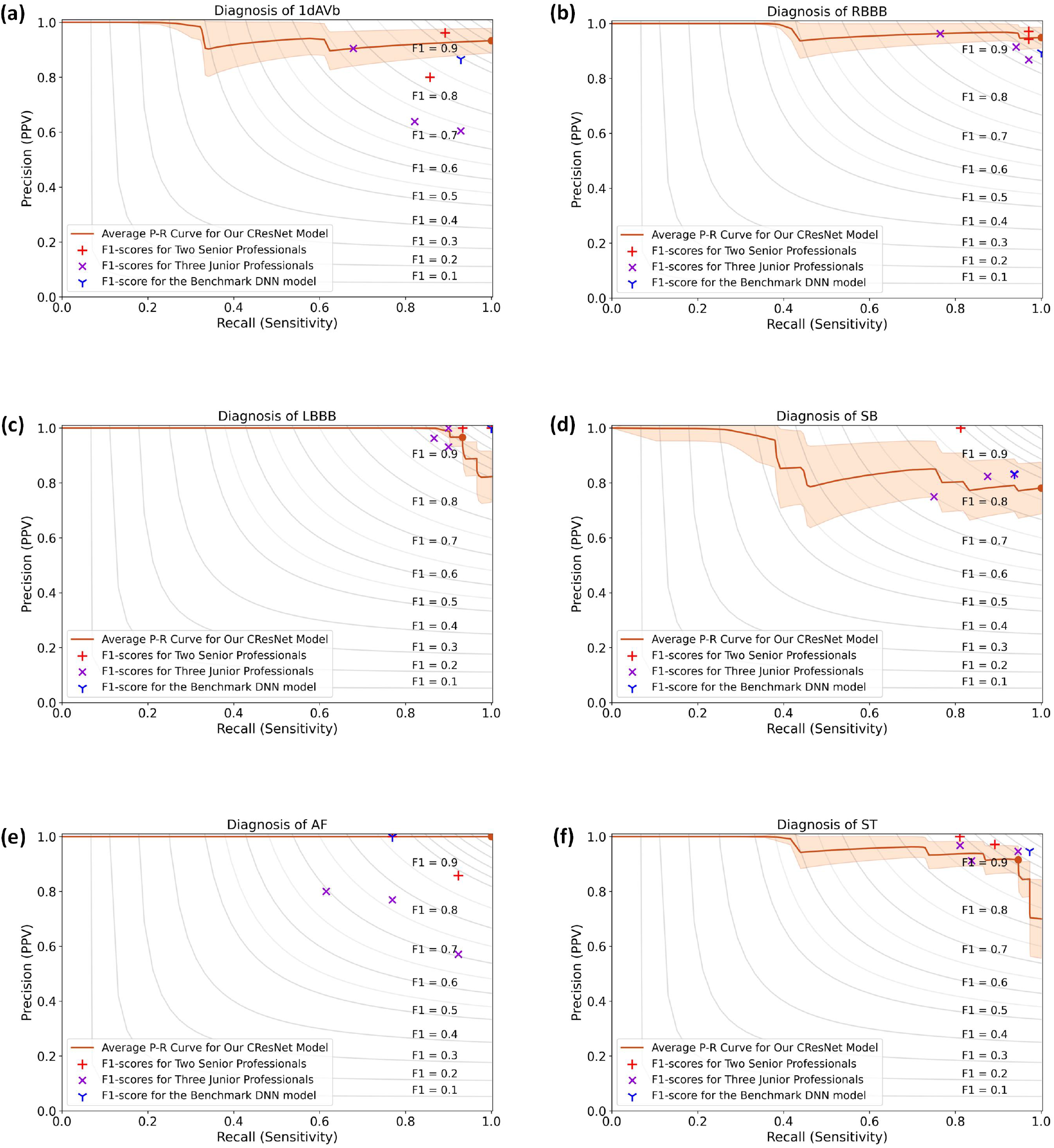
Performance comparison for the diagnosis of abnormalities, including **(a)** 1dAVb, **(b)** RBBB, **(c)** LBBB, **(d)** SB, **(e)** AF, and **(f)** ST. This figure shows the precision-recall (P-R) curves for the performance of the CResNet model, evaluation results from five cardiology professionals, and the result of the benchmark DNN model [38]. The solid lines are the average P-R curves for the diagnosis of arrhythmias, and the shading areas represent standard deviations obtained by the bootstrap method. The brown dots correspond to the F1-scores for the CResNet model, the red ‘+’ symbols are used to denote F1-scores for the two senior professionals, the purple ‘X’ for the three junior professionals, and the blue ‘Y’ for the benchmark DNN model. The contour plots show the iso-F1 curves with a constant value for each curve, and a point closes to the ideal score of ‘1’ in the top-right corner indicating a higher F1-score.

Among these abnormalities, the diagnosis of AF particularly has important medical implications, because it is a leading cardiac cause of stroke, heart failure, and mortality [39]. However, it is challenging to obtain a definitive diagnosis of AF with ECG recordings [30], which is also indicated by the evaluation results as presented in Table 1A. It can be seen from the table that among all the five professionals, the highest F1-score for the diagnosis of AF is 0.889 (95% CI, 0.737-1.000); and the benchmark model from the literature also has moderate performance with a score of 0.870 (95% CI, 0.667-1.000) [38]. In contrast, our developed CResNet model successfully identifies all AF in the dataset.

Next, we interpret how the decisions that have been made by the CResNet model to diagnose ECG abnormalities. Figure 3(a) shows a standard 12-lead ECG recording with AF identified in the test. Five cardiology professionals evaluated the test, and only one of the senior cardiologists and the emergency resident successfully diagnosed the AF; while the other senior and two junior professionals failed to diagnose it. Using our developed CResNet model, the diagnosis of AF with the ECG recording has a prediction probability of 0.961. To interpret the diagnosis that has been identified by the CResNet model, we calculate the heatmap for each of the 12 ECG leads, and highlight the salient information that has been used for decision making. In Figure 3(a), the different colours indicate weights of data points in the ECG recording, e.g., red colour for important information with a high weight, and blue for less important data with a low weight. It can be seen from Figure 3(a) that the CResNet model uses salient information in the DII and V1 leads for the diagnosis of AF, and has the most important features with red colour in the DII lead.

**Figure 3:**
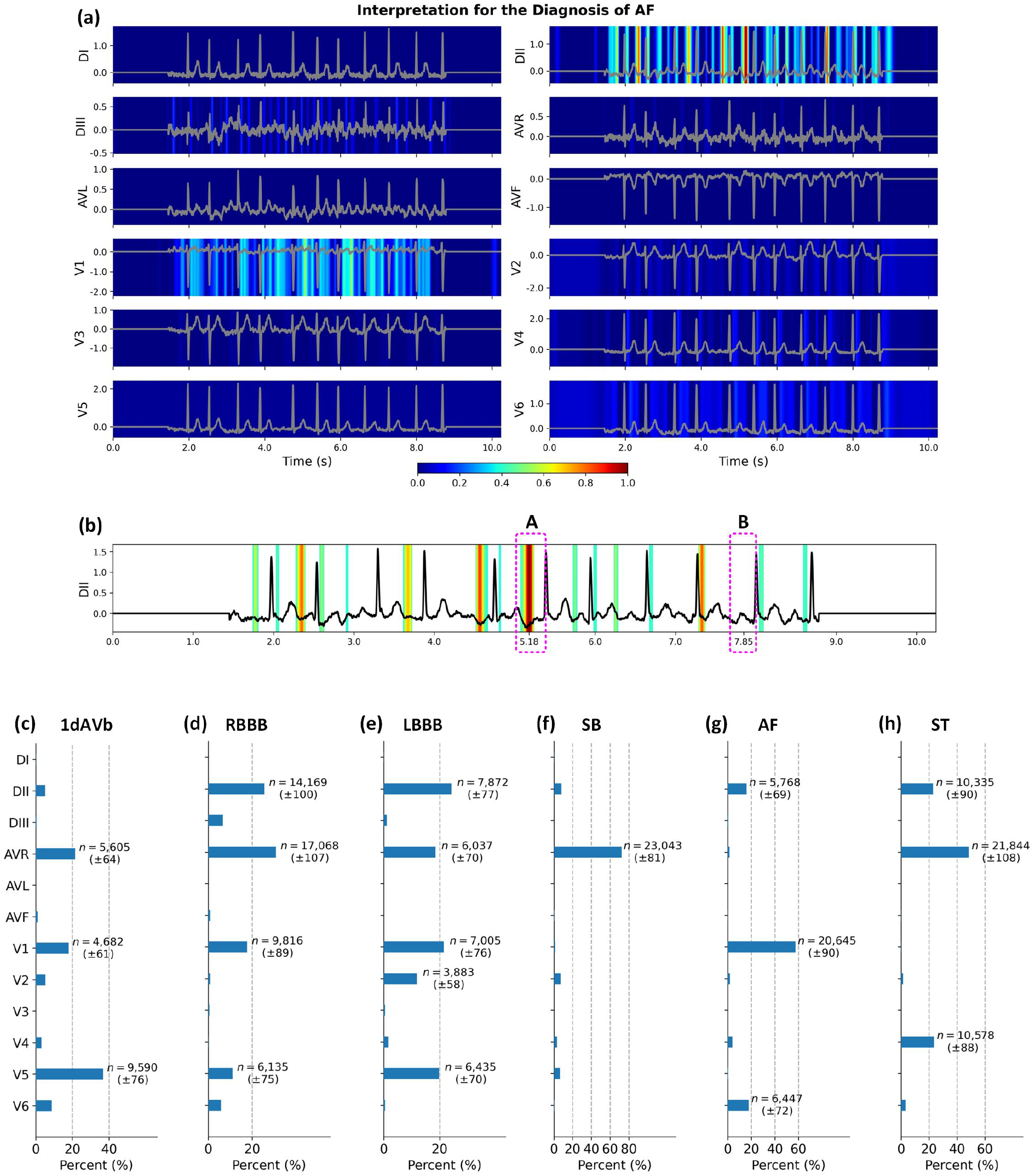
Interpretation for the diagnosis of AF and distributions of dominant ECG leads. **(a)** The original calculated heatmaps for the diagnosis of AF using 12 ECG leads, with colour bar ranging from blue to red indicating the increasing weights of data importance. **(b)** The refined view of the DII lead in **(a)** by removing background colours with values less than 0.4. Segments *A* and *B* show the inconsistent morphologies in the locations of P waves in the DII lead. We show distributions of dominant ECG leads for the diagnosis of **(c)** 1dAVb, **(d)** RBBB, **(e)** LBBB, **(f)** SB, **(g)** AF, and **(h)** ST. We annotate the number of occurrences when the dominant lead accounts for more than 10% of all the 12 ECG leads. The number of occurrences is presented as mean and standard deviation calculated by bootstrap method.

Notably, the hallmark of AF is the absence of P waves in an ECG recording [30]. However, artifacts or fibrillatory waves can mimic P waves and lead to misdiagnosis [40, 41]. Figure 3(b) shows the refined view of the DII lead with background colour removed, which demonstrates the ECG morphology and salient features that have been used for the diagnosis. It can be seen from Figure 3(b) that the P wave is absent in some areas of the ECG morphology, e.g., segment *A* (around 5.18s); and there are also waves clearly presented in some areas, e.g., segment *B* (around 7.85s). The inconsistent morphologies in the locations of P waves challenge the diagnosis of AF using the ECG recording. Our developed CResNet model is very flexible in the recognition of P waves, and it highlights important information in segment *A* rather than segment *B*, which is consistent with the existing diagnostic criteria [40, 41]. As well as identifying the absence of P waves, the CResNet model also recognises S waves as salient features in the DII lead, and other features in the V1 lead. With combining salient information from different leads in the ECG recording, the CResNet model makes a comprehensive decision with the prediction probability of 0.961 for the diagnosis of AF.

Other than the interpretation for the diagnosis of AF, we present salient features that are used to diagnose other types of ECG abnormalities in Extended Figures S4-S8. The results demonstrate that the interpretation that has been made by the CResNet model matches well with existing knowledge, but also provides new implications with the identified salient features. For example, as shown in Extended Figure S4, the CResNet identifies the absence of Q waves, notched R waves, and T wave inversion in the V6 lead for the diagnosis of LBBB; Furthermore, it highlights the absence of Q waves and T wave inversion in the DII lead even with higher weights. Combining salient features in the 12 ECG leads, the CResNet model diagnoses the LBBB with a probability of 0.948. In another example as illustrated in Extended Figure S7, the CResNet model identifies the U waves in the AVR lead, and uses them as important information for the diagnosis of SB. This is consistent with previous observations of prominent U waves in the ECG recording [42]; Apart from identifying U waves in the AVR lead, the CResNet also identifies the downslopes of T waves in DII lead as important information, and the model has a probability of 0.932 to diagnose the SB with combining salient information in the ECG recording.

In a further step, because ECG abnormalities have varied morphologies, we present the statistical results of dominant leads that are derived from the salient information. First, we filter ECG recordings in the whole dataset with prediction probabilities higher than 0.8, which indicates the CResNet model having confident outputs for the diagnosis of abnormalities. Then, we sum the values of the heatmap for each lead, and identify the dominant lead with the highest value for the ECG recording. To show distributions of the identified dominant leads, we calculate their occurrences and the percentages among all the 12 ECG leads, and the results can be found in Figures 3(c)-(h). It can be seen from Figures 3(c)-(h) that the six types of ECG abnormalities have varied distributions of dominant leads. The 1dAVb has AVR, V1, and V5 as dominant leads; both the RBBB and LBBB have dominant DII, AVR, V1, and V5 leads; the SB has a prominent AVR lead; the AF has three dominant leads of DII, V1 and V6; and the ST has DII, AVR and V4 as dominant leads.

Next, we investigate the effectiveness of the identified dominant leads on the diagnosis of ECG abnormalities. As shown in Figures 3(c)-(h), the AVR and V1 leads are two representative leads for the ECG abnormalities. We therefore use the two leads to train the CResNet model, and test the performance on the holdout dataset. Table 1B shows the results of the diagnosis using the AVR and V1 leads. It can be seen from the table that the CResNet model achieves an AUC score of 0.990 (95% CI, 0.982-0.995) and an F1-score of 0.879 (95% CI, 0.834-0.919) using the two dominant leads, which is comparable to the best performance of the three junior professionals (*p* = 0.505). In addition to the dominant AVR and V1 leads, the DII lead is also representative for all types of ECG abnormalities. With including the DII lead, the CResNet model achieves an AUC score of 0.995 (95% CI, 0.992-0.997) and an F1-score of 0.903 (95% CI, 0.868-0.935). In particular, using the DII, AVR, and V1 leads, the model has an F1-score of 0.917 (95% CI, 0.750-1.000) for the diagnosis of AF, and 0.923 (95% CI, 0.844-976) for the diagnosis of ST, which is higher than the scores of 0.889 (95% CI, 0.727-1.000) and 0.880 (95% CI, 0.786-0.956) using the AVR and V1 leads.

**Table 1B:** Performance comparison for the diagnosis of abnormalities using ECG recordings with dominant leads.

|  | Dominant AVR and V1 Leads |  |  | Dominant DII, AVR, and V1 Leads |  |  |
| --- | --- | --- | --- | --- | --- | --- |
|  | Precision<br>(95% CI) | AUC<br>(95% CI) | F1-score<br>(95% CI) | Precision<br>(95% CI) | AUC<br>(95% CI) | F1-score<br>(95% CI) |
| 1dAVb | 0.870<br>(0.714 - 1.000) | 0.991<br>(0.982 - 0.998) | 0.784<br>(0.632 - 0.898) | 0.952<br>(0.842 - 1.000) | 0.992<br>(0.982 - 0.998) | 0.816<br>(0.667 - 0.927) |
| RBBB | 0.935<br>(0.833 - 1.000) | 0.996<br>(0.992 - 0.999) | 0.892<br>(0.800 - 0.964) | 0.909<br>(0.800 - 1.000) | 0.997<br>(0.993 - 0.999) | 0.896<br>(0.807 - 0.966) |
| LBBB | 0.966<br>(0.889 - 1.000) | 0.978<br>(0.931 - 1.000) | 0.949<br>(0.875 - 1.000) | 1.000<br>(1.000 - 1.000) | 0.993<br>(0.982 - 1.000) | 0.947<br>(0.880 - 1.000) |
| SB | 0.762<br>(0.565 - 0.947) | 0.998<br>(0.995 - 1.000) | 0.865<br>(0.722 - 0.973) | 0.842<br>(0.647 - 1.000) | 0.999<br>(0.996 - 1.000) | 0.914<br>(0.786 - 1.000) |
| AF | 0.857<br>(0.636 - 1.000) | 0.998<br>(0.993 - 1.000) | 0.889<br>(0.727 - 1.000) | 1.000<br>(1.000 - 1.000) | 0.997<br>(0.992 - 1.000) | 0.917<br>(0.750 - 1.000) |
| ST | 0.868<br>(0.750 - 0.972) | 0.997<br>(0.993 - 0.999) | 0.880<br>(0.786 - 0.956) | 0.878<br>(0.757 - 0.973) | 0.998<br>(0.995 - 1.000) | 0.923<br>(0.844 - 0.976) |
| Micro_avg | 0.885<br>(0.830 - 0.937) | 0.990<br>(0.982 - 0.995) | **0.879**<br>**(0.834 - 0.919)** | 0.921<br>(0.875 - 0.962) | 0.995<br>(0.992 - 0.997) | **0.903**<br>**(0.868 - 0.935)** |

\*The term of ‘Dominant AVR and V1 Leads’ indicates that the model has inputs with only two ECG leads, e.g., AVR and V1 leads, rather than using 12 ECG leads.

Additionally, we validate our developed CResNet model on an external dataset, which is retrieved from the PhysioNet/CinC Challenge 2017 [43]. The dataset consists of short single-lead ECG recordings that have been annotated with four classes, i.e., normal sinus rhythm, atrial fibrillation, other alternative rhythms, and noise. We train the CResNet model using ECG recordings (*n*_ECGs_ = 8,528) in the training dataset, and test the model on the holdout validation dataset. Extended Table S4 presents the model performance for classifying the four types of ECG recordings. It can be seen from the table that the CResNet model has a micro average F1-score of 0.884 on the validation dataset. In particular, the model has an F1-score of 0.929 on the diagnosis of AF, and a score of 0.921 on detecting noise signals; Given the widespread noises in ECG recordings, the results indicate robustness of our model for the diagnosis of heart rhythm abnormalities.

### 2.3 Identification and Interpretation of Genders

In the second task, as demonstrated in Figure 4(a), our developed CResNet model has an AUC score of 0.964 (95% CI, 0.963-0.965) on gender identification for individual subjects in the holdout testing dataset (*n*_Subjects_ = 155,435). Because features presented in ECG recordings may change over time due to normal ageing [44], we therefore investigate the model performance in different age groups [45], i.e., young-age (years (*yr*) *<* 45, *n*_Subjects_ = 54,341), middle-age (45 ≤ *yr <* 75, *n*_Subjects_ = 84,640), and old-age (*yr* ≥ 75, *n*_Subjects_ = 16,454). It can be seen from Figure 4(a) that the CResNet model has an AUC score of 0.979 (95% CI, 0.977-0.980) on identifying genders for the young-age group, which is higher than the AUC score of 0.959 (95% CI, 0.958-0.961) for middle-age group, and 0.914 (95% CI, 0.909-0.918) for old-age group, suggesting the effect of ageing on the gender identification (*p <* 0.01) using standard 12-lead ECG recordings.

**Figure 4:**
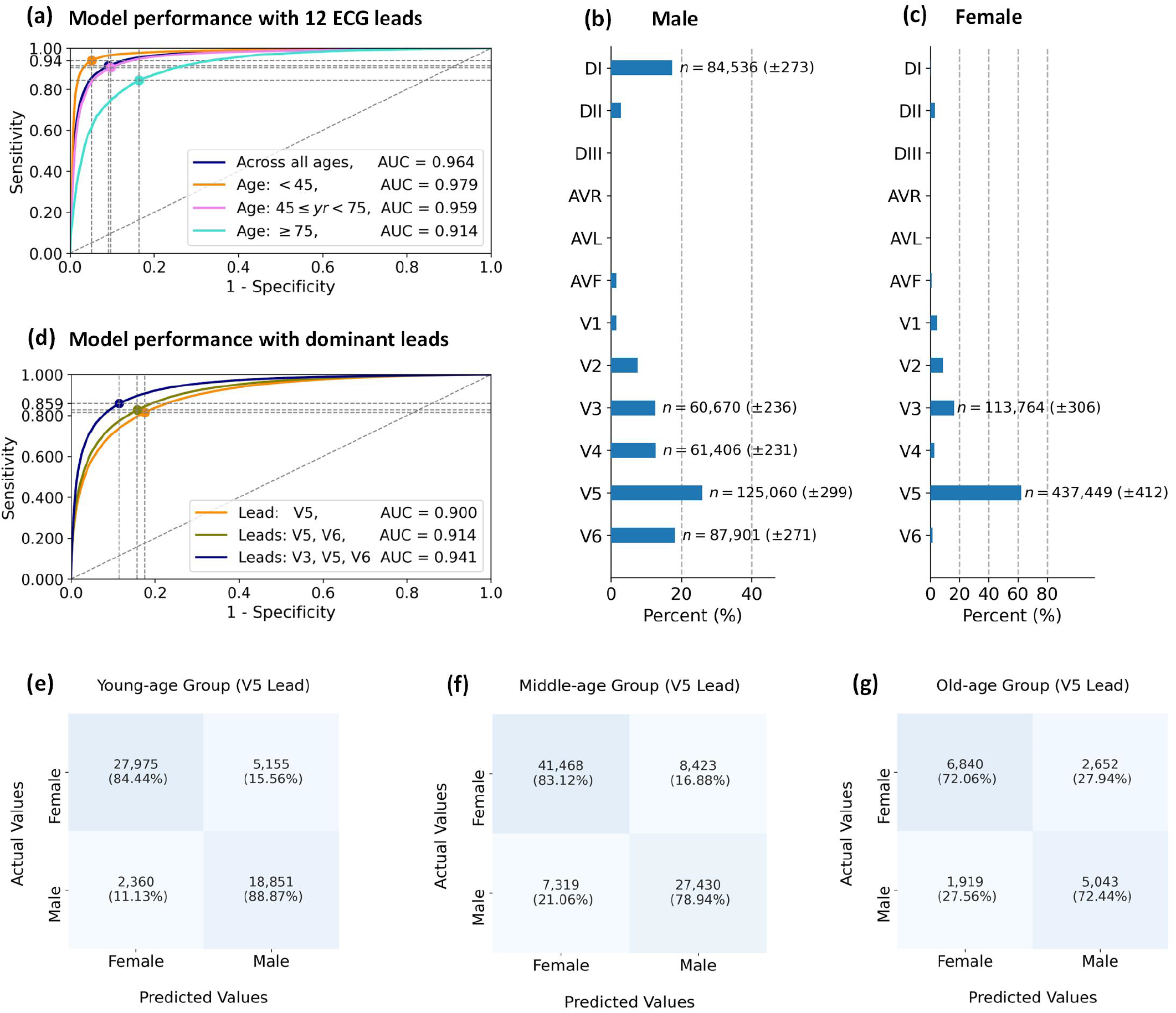
Model performance and lead importance for gender identification using our proposed CResNet model. **(a)** Performance comparison of the CResNet model for gender identification using 12-lead ECG recordings in different age groups. **(b)** Distributions of dominant leads for identifying male subjects. **(c)** Distribution of dominant leads for identifying female subjects. **(d)** Performance comparison between different dominant ECG leads. We demonstrate confusion matrices for gender identification using the dominant V5 lead in different age groups, including **(e)** the young-age group (*yr <* 45), **(f)** the middle-age group (45 ≤ *yr <* 75), and **(g)** the old-age group (*yr* ≥ 75). In each of the receiver operating characteristic (ROC) curves, the dot point indicates the optimal cut-off point for the sensitivity and specificity calculated by the G-mean method.

To show the interpretation of gender identification, we visualise the salient features in ECG recordings for identifying female in Extended Figure S10, and male in Extended Figure S11. It can be seen from Figure S10 that the model mainly uses salient information from the DII, V1, and V5 leads for identifying the female subject, which has a prediction probability of 0.971; For identifying the male subject, the model uses the DI, V4, V5, and V6 leads and has a probability of 0.981. We then use the post-hoc method as presented in the first task to analyse the distribution of dominant leads for gender identification. Figures 4(b) and (c) present distributions of dominant leads for identifying male and female subjects separately. The detailed distributions of dominant leads in terms of age differences, i.e., young-age, middle-age, and old-age, can be found in Extended Figure S12. It can be seen from Figures 4(b) and (c) that V5 is the mostly used lead for gender identification by the CResNet model, which is a dominant lead for identifying male subjects (*n*_Male_ = 125,060 ± 299) and female subjects (*n*_Female_ = 437,449 ± 412). Other than the V5 lead, the V3 lead also appears as a dominant lead for identifying male subjects (*n*_Male_ = 60,670 ± 236) and female subjects (*n*_Female_ = 113,764 ± 306).

Next, we investigate the model performance of gender identification using the identified dominant leads, and the comparison results are presented in Figure 4(d). As the V5 lead is dominant for both male and female subjects, we first only use the V5 lead to identify gender, and the CResNet model obtains an AUC score of 0.900 (95% CI, 0.899-0.902). We show confusion matrices of gender identification in different age groups using the dominant V5 lead in Figures 4(e)-(g), and it can be seen that the CResNet model has the highest performance in the young-age group (*p <* 0.01), with an accuracy of 84.44% for identifying female subjects and 88.87% for identifying male subjects. Apart from the V5 lead, we note that the V6 is identified as a dominant lead for identifying males, but less important for females as shown in Figure 4(c). Therefore, we combine V6 with V5 to implement the gender identification, and the model has a slightly higher AUC score of 0.914 (95% CI, 0.913-0.915) with the two dominant leads (*p <* 0.01). In a further step, we include the V3 lead to generate a new combination of three dominant leads for gender identification, and the AUC score increases from 0.914 (95% CI, 0.913-0.915) using the V5 and V6 leads to 0.941 (95% CI, 0.940-0.943) using the V3, V5, and V6 leads, indicating the importance of the V3 lead for gender identification (*p <* 0.01).

In addition, we present comprehensive comparisons of model performance using different combinations of dominant ECG leads for gender identification in Extended Figures S14-S16 and Tables S5-S7. The results show that using the DI, V3, and V5 leads, the CResNet model has the highest performance (*p <* 0.01) with an AUC score of 0.970 (95% CI, 0.969-0.972) and a diagnostic odds ratio (DOR) of 145.891 (95% CI, 139.089-156.331) for gender identification in the young-age group (Extended Figure S14). We note that all models have lower performance on identifying genders in the old-age group than in the young-age group (*p <* 0.01), with the highest AUC score of 0.885 (95% CI, 0.880-0.890) in the old-age group using the DI, V3, and V5 leads. The comparison results of model performance suggest the effectiveness of our identified dominant ECG leads for gender identification.

### 2.4 Screening and Interpretation of Hypertension

In parallel with the previous two tasks, we implement the third task of hypertension screening using our developed CResNet model, and the results of model performance are presented in Figure 5(a) and (b). It can be seen from Figure 5(a) that the CResNet model achieves an AUC score of 0.839 (95% CI, 0.837-0.841) and a diagnostic odds ratio (DOR) of 12.101 (95% CI, 11.794-12.447) in screening subjects with hypertension in the testing dataset (hypertension: 31.65%, *n*_Hypertension_ = 49,202). Considering the effects of age and gender on the prevalence of hypertension [44], we investigate the model performance of hypertension screening in different populations. It can be seen from Figure 5(a) that the model achieves an AUC score of 0.849 (95% CI, 0.847-0.852) for hypertension screening in the female group, which is slightly higher than the AUC score of 0.823 (95% CI, 0.820-0.827) in the male group (*p* = 0.011). In terms of age differences, as shown in Figures 5(b) and (c) that the model has the highest performance in the old-age female group (*p <* 0.01), with an AUC score of 0.829 (95% CI, 0.822-0.836) and the DOR of 18.172 (95% CI, 16.516-20.576).

**Figure 5:**
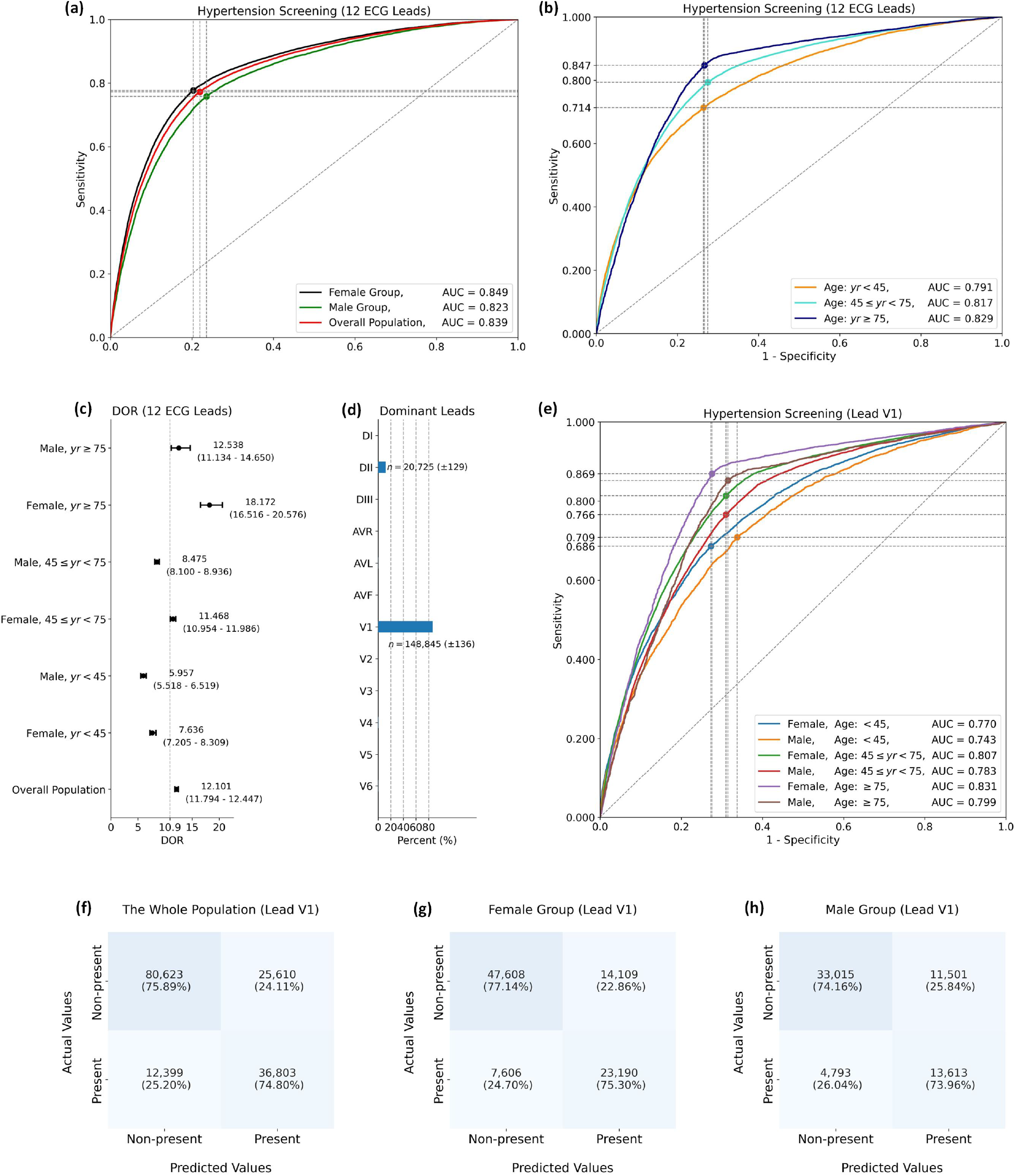
Model performance and lead importance for hypertension screening using our developed CResNet model. **(a)** Performance comparison of the CResNet model for hypertension screening using 12-lead ECGs in terms of gender differences. **(b)** Performance comparison in terms of age differences using 12-lead ECGs. **(c)** Diagnostic odds ratios (DOR) with 95% CI for hypertension screening in different populations. **(d)** Distributions of the dominant ECG leads (mean ± standard deviation). **(e)** Performance comparison of hypertension screening using the dominant V1 lead. We demonstrate confusion matrices for hypertension screening using the dominant V1 lead in different population groups, including **(f)** the whole population, **(g)** the female group, and **(h)** the male group. The confidence interval and standard deviation are calculated by bootstrap method.

To show the interpretation of hypertension screening, we visualise the salient features in the 12 ECG leads, which have been used to make decision by the CResNet model. As an illustration in Extended Figure S17, the CResNet model mostly uses the DII, AVL, and V1 leads to screen hypertension, with particular focuses on T waves in the DII and V1 leads. Next, we use post-hoc analysis to identify dominant ECG leads from the salient features, and investigate their performance on hypertension screening.

It can be seen from the distribution in Figure 5(d) that the CResNet model identifies the DII and V1 as dominant ECG leads. In particular, the V1 lead accounts for more than 80% of the occurrences among the 12 ECG leads, which is used to screen hypertension for *n*_Hypertension_ = 148,845 ± 136 subjects; Other than the V1 lead, the DII lead is also identified as a dominant lead to screen hypertension for *n*_Hypertension_ = 20,725 ± 129 subjects.

We therefore use the dominant V1 lead to screen hypertension for individual subjects, and it can be seen from Figure 5(e) that the model obtains an AUC score of 0.831 (95% CI, 0.823-0.840) in the old-age female group, which is a similar result to the model performance of 0.839 (95% CI, 0.837-0.841) using 12 ECG leads; and as shown in Figures 5(f)-(h) that the CResNet model has an accuracy of 74.80% on screening hypertension in the whole population using the V1 lead, and it has a higher accuracy of 75.30% in the female group than in the male group. In a further step, we use two ECG leads to screen hypertension with including the additional DII lead, which achieves the highest AUC score of 0.835 (95% CI, 0.827-0.844) in the old-age female group (Extended Figure S19). As shown in Extended Figures S21-S23 and Tables S9-S11, we present the detailed comparison of hypertension screening using dominant ECG leads and 12 ECG leads in terms of age and gender differences, and the results suggest effectiveness of our identified dominant leads for hypertension screening in different population groups.

### 3. Discussion

In this study, we developed and validated a novel ‘end-to-end’ DNN model with state-of-the-art performance on medical diagnosis, and in particular it has the ability to interpret the ECG recording that has been used to make the decision by the AI model. Our work builds on the recent research in literature that shows deep learning can be used to diagnose ECG abnormalities with cardiologist-level performance [26, 30, 38]. However, these models are seen as black boxes, which primarily focus on improving the accuracy of arrhythmia detection. It is therefore difficult to explain how the methods underlying the ECG morphologies have been used for the diagnosis. Given the rising demand for explanatory AI models from clinicians and government regulators [1, 10, 12, 13, 14], our study presents a substantial response towards developing an interpretable deep learning model for healthcare.

There are several initial studies in the literature that show computerised interpretation of ECG recordings [5, 46, 47, 48]. As a representative example, the study in [5] developed a DNN model to predict 1-year all-cause mortality using 12-lead ECG recordings. The research in [5] obtained attractive results with an AUC score of 0.88, and derived prognostic information for mortality prediction, which had important implications in medical practice; In a further step, the research interpreted ECG recordings and indicated the relationship between ST segments and mortality prediction [5]. However, the interpretation of lead-specific ECG recordings is still challenging for several reasons. First, the research used multiple leads together as model inputs, the learned saliency map was therefore shared by several leads rather than the accurate weight for a specific ECG lead. Although a guided-backpropagation technique [20] was used to derive lead-wise weight, the generated heatmap for the ECG recording was discrete. For example, adjacent data in the ECG morphology was highlighted as disconnected points in the heatmap, which makes it difficult to understand the visualised salient features; Second, it is understood that ECG morphology may change over time. As demonstrated in Figure 3(b) in our study, some morphologies, e.g., segments in the location of P waves, may be not constant in the ECG recording. It therefore requires the interpretation of ECGs to be flexible and accurate over time. While the study in [5] demonstrated the interpretation for ECG data segments with a short duration, i.e., 0.6s, and it is not clear how the interpretability changes over time when standard or longer duration ECG recordings are used. Third, from a clinical perspective, it is difficult to verify the interpretation of specific morphologies for the prediction of mortality or rare diseases.

Our developed interpretable DNN model was first implemented on the diagnosis of arrhythmias, which has been well studied in the literature and existing knowledge can be used to validate our findings. A recent study showed the importance of DII lead in the diagnosis of abnormalities, which was used to classify twelve rhythm classes [26]; For the diagnosis of ventricular arrhythmias, the V1 lead was observed having dominant waves [49]; In addition, lateral leads (e.g., lead V5) were considered to be important in the diagnosis of bundle branch blocks [50, 51]. Consistent with prior knowledge of dominant ECG leads for the diagnosis of arrhythmias, our findings also provide new insights; For example, the diagnosis of SB and ST primarily focuses on the checking of a patient’s heart rate by cardiologists, whereas our CResNet model highlights the importance of U waves for the identification of SB. Notably, prominent U waves were also reported in asymptomatic SB in literature [42]. However, U waves are usually difficult to be measured in manual review due to their low amplitudes [52], which in contrast suggests the advantages of our model for computerised ECG interpretation. In particular, we highlight the benefits from our proposed isolation-integration strategy, which allows the model to precisely calculate the importance of each ECG lead, and therefore enables to identify lead-specific and disease-specific features for the diagnosis of different arrhythmias. After validating our developed interpretable model in the first task of the diagnosis of ECG abnormalities, we then extended the study in a wider context of medical diagnosis, i.e., the second task of gender classification and the third task of hypertension screening; Again, a previous study indicated the importance of V5 lead for gender identification [48], confirming the findings of our identified dominant leads. We note that previous studies indicated that high blood pressure is in association with an increased risk of AF [53], this could explain the AF and hypertension having similar dominant ECG leads in our study, i.e., the DII and V1 leads. Finally, we highlight that our interpretation was implemented on standard 12-lead ECG recordings, instead of a specific lead or limited sampling duration; this moves research efforts a step closer towards the application of interpretable AI algorithms in medical practice.

From a clinical perspective, the interpretation of ECG recordings is critical to understand and diagnose cardiovascular diseases. Our developed DNN model can potentially augment the current clinical workflow in several ways. First, rather than developing a stand-alone computerised method for automated ECG diagnosis, our CResNet model presents a paradigm shift by producing visually salient features for the interpretation of ECG recordings, which allows practitioners to understand the decision that has been made by the AI model, and therefore reduce the risk of misdiagnosis. Second, benefiting from the visually salient features and the identified dominant ECG leads, our developed model has the potential to facilitate the discovery of new biomarkers, particularly in areas where expert knowledge is not readily available, i.e., hypertension screening using ECG recordings. Third, even in well-established area, e.g., diagnosis of arrhythmias, our developed DNN model can provide new insights for the interpretation of ECG morphologies; this enables us to promote further understanding of cardiovascular systems. Notably, the model developed in this study does not involve any prior domain knowledge, i.e., cardiovascular medicine, but instead allows automated learning of salient features in data measurements that are collected from physically isolated sensors. It is therefore that our CResNet model could potentially be used in other scenarios apart from the medical tasks performed in this study. For example, it could be applied to identify the importance of different channels for electroencephalography (EEG) signals, which allows further understanding of brain activities.

We note that the standard 12-lead ECG test is a widely used non-invasive screening tool in healthcare, and recent advances in ECG technologies have enabled the development of small, low-cost, and easy-to-use wearable devices in limited-resources or home settings [54], which typically use a subset of the standard 12 ECG leads for remote monitoring. However, as highlighted in the recent PhysioNet/Computing Challenge [55], there is limited research to demonstrate that reduced-lead ECGs can capture the wide range of diagnostic information achieved by the 12-lead ECGs. The study in this paper provides substantial evidence to show that our developed AI model enables to automatically identify important ECG leads for various medical tasks other than the diagnosis of cardiac abnormalities. By undertaking extensive comparison studies between our identified dominant ECG leads and the standard 12 ECG leads, we show that our identified reduced-lead ECGs can achieve comparable performance with the standard 12-lead ECGs for lead-specific and disease-specific diagnosis, which can meaningfully contribute to the development of reduced-lead wearable devices for cardiac monitoring. In particular, our proposed CResNet demonstrates the effectiveness of hypertension screening using reduced-lead ECGs, indicating the potential applications of our developed model for cuffless blood pressure estimation/monitoring as a future direction.

Our work is perhaps best understood in the context of its limitations. We note that there are a broad range of heart arrhythmias, and the current study tested the interpretation with a limited category of abnormalities. However, the diagnosis of different types of arrhythmias has a similar procedure in the analysis of ECG morphologies, and our model learns salient features in ECG recordings with no assumption of a specific type of abnormalities. It is therefore that the CResNet model could be possibly extended to study other abnormalities when the datasets are available. We note that in order to perform accurate visualisation of salient features, it is vital to recognise that ECG morphology varies between subjects, and that, in some cases, indeed there are exceptions that it might not be possible to derive accurate diagnostic information from the ECG morphology features. To select representative features that are used for the diagnosis, we first use post-hoc analysis to calculate prediction probabilities for all the ECG recordings, and then screen them with a threshold of probability larger than 0.8, which represents an acceptable level of confidence that the model can accurately make the prediction using the features identified in the ECG test. In a further step, we present comprehensive results of the identified features across a large cohort of the population by providing statistical distributions of dominant leads that are derived from the salient features. We also note that in addition to hypertension screening using ECGs as presented in this study, there are many other life-threatening diseases to which the model could be applied, such as myocardial infarction, ventricular tachycardia or other cardiac conditions. Our future research will use the developed model to perform interpretation for other medical tasks in a wider diagnostic context. Additionally, it is well accepted that a DNN model is complex and highly nonlinear, and it is nearly impossible to explain the whole inference process of the decision making [1]. The current study leverages the advances of techniques from image visualisation to derive salient features for automated ECG interpretation; the relationship between these different features and how they interact and lead to a comprehensive diagnosis are potentially valuable areas for more detailed investigation in future.

In conclusion, we demonstrate an end-to-end deep learning model with cardiologist-level performance outperforming the state-of-the-art in medical diagnosis, and more importantly, the model provides an interpretation of ECG recordings that have been used for decision making. Using a sufficiently large dataset (2.3 million ECGs collected from 1.6 million subjects), we validated the performance of our model on three independent medical tasks, such as arrhythmia diagnosis, gender identification, and hypertension screening. We showed that our model provides substantial advantages to promote accurate diagnosis by producing visually salient features, and that in particular, it has potential to enhance the understanding of diagnostic decisions for different diseases, and to discover novel patient-relevant information from clinical data measurements.

## Data Availability

All data produced in the present work are contained in the manuscript.

## 4 Methods

### 4.1 Data Acquisition and Annotation

The present study uses a dataset consisting of standard 12-lead ECG recordings collected by the Telehealth Network of Minas Gerais (TNMG), a public healthcare system to provide tele-consultation and tele-diagnosis for 811 municipalities in the state of Minas Gerais, Brazil [38, 56]. The ECG recordings were mostly collected in primary care facilities during clinic visits between 2010 and 2016, which were performed either using the tele-electrocardiogram machine of model TEB (Tecnologia Eletrônica Brasileira, São Paulo, Brazil), or the ErgoPC 13 (Micromed Biotecnologia, Brasilia, Brazil). This study complies with all relevant ethical regulations, and the Research Ethics Committee of the Universidade Federal de Minas Gerais (Protocol 49368496317.7.0000.5149) gave the ethical approval.

The ECG tests were recorded for a duration of 7 to 10 seconds with sampling frequencies ranging from 300 to 600 Hz. To ensure consistency of the data format, the recordings were resampled with 400 Hz, and then zero-padded to the length of 4,096 data points. The rescaled ECG recordings were stored in a structured database, namely the Clinical Outcomes in Digital Electrocardiology (CODE). A cohort of 2,322,513 ECG recordings were retrieved from the CODE dataset. We excluded lowquality ECGs (*n*_ECGs_ = 6,731) that had zero values for more than 80% of the data points, and used a total of 2,315,782 ECG recordings for the current study.

We obtained electronic health records for subjects in the CODE dataset by performing a link matching between the ECG tests and the national mortality information system, using a standard probabilistic linkage method (FRIL: Fine-grained record integration and linkage software, v.2.1.5, Atlanta, GA) [56, 57]. Hypertension in the health records was defined as a systolic blood pressure ≥ 140 mm Hg, or diastolic blood pressure ≥ 90 mm Hg, or self-declared use of anti-hypertensive medication. The data were anonymised after the linkage matching.

Annotation of ECG recordings in the CODE dataset was performed by both trained professionals and computerised software [58] using the following procedures, (*i*) the sampled ECG recordings were first sent by internet to central servers, and a team of trained professionals used standardised criteria to generate free-text ECG reports [59], which were digitally recognised by a hierarchical free-text machine learning method [60]. The ECG reports were periodically audited by professionals to recognise medical errors and discordant interpretations; (*ii*) The Glasgow 12-lead ECG analysis program was used to analyse the ECG recordings [61], and generate the diagnosis results of the Glasgow Diagnostic Statements and Minnesota Code [61, 62]; (*iii*) The presence of a specific ECG abnormality was automatically considered when there was an agreement between the cardiologist report and the computerised diagnosis result. A manual review was performed when the two sources of diagnosis disagreed [56].

The holdout testing dataset for model evaluation was independently and rigorously reviewed by two certified cardiologists, and the data label was obtained when annotations from the two professionals were matched; Where annotations did not match, a specialist was introduced to decide the diagnosis. We present the evaluation results of the two senior professionals in Table 1A. We also calculate the Cohen’s kappa coefficient of the evaluation results from the two senior professionals [38]; values are 0.741 for 1dAVb, 0.955 for RBBB, 0.964 for LBBB, 0.844 for SB, 0.831 for AF, and 0.902 for ST. These values demonstrate the inter-rater agreement for the two professionals, and we therefore use these evaluation results as the data labels. The testing dataset was also reviewed by three groups of junior cardiology professionals, i.e., two 4^*th*^ year cardiology residents, two 3^*rd*^ year emergency residents, and two 5^*th*^ year medical students. To reduce the bias of ECG evaluation, the two professionals in each of the three groups were asked to annotate half of the testing dataset, and the concatenated performance scores were obtained for the three groups.

### 4.2 Model Development

Our method developed in this study consists of two modules; the first one is a DNN model that is used to make inference for the diagnosis, and architecture of the DNN model is illustrated in Extended Figures S2. The second module is an interpretation model to produce salient features for ECG recordings, and the flowchart is described in Extended Figures S3. As the DNN model is developed using the mechanism of the interpretation model, we next introduce the principles of developing an interpretation model with refined resolution, and then describe the details of developing the DNN model.

#### 4.2.1 Principles for Designing the Interpretation Model

The interpretation model is developed using a refined gradient-weighted class activation mapping (Grad-CAM) module. The Grad-CAM assumes that the last convolutional (Conv) layer in a deep learning model represents higher-level visual content of the input data [20, 63]. Then, the model calculates the gradient information with respect to the last Conv layer, and uses it to represent the importance of each kernel for the decision making.

Formally, for the input data **X** and corresponding label *y* ∈ 𝒩^*c*^, a deep learning model with convolutional neural networks builds mapping for the input data and output label, *f* : **X** → *y*. The Grad-CAM model first computes the gradient score for class *y*^*c*^ with respect to the feature map **W** in the last Conv layer [20, 63],

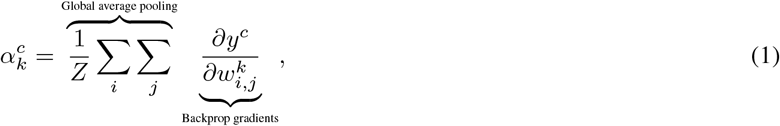

where, 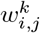 is the element of feature map **W** in the last Conv layer, 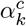 is the calculated weight, which is used to weight the importance of the *k*^*th*^ kernel in the feature map.

Next, a coarse localisation heatmap can be obtained by a weighted combination of feature maps, and it is followed by an activation function,

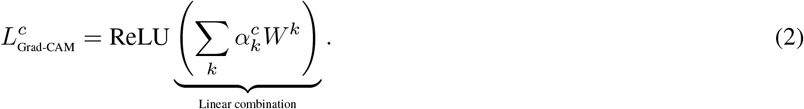

where, ReLu(·) is the rectified linear unit function [64], which is used to find a positive influence on the class of interests; matrix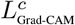 is the calculated heatmap for the *y*^*c*^ class, and the calculated heatmap has the same dimension of the kernel size.

The Grad-CAM has been widely used for the interpretation of image data, however, as demonstrated in previous research, the target objects detected by the model include much irrelevant information [20, 21]. This is mostly due to the following reasons: (*i*)*Dimension alignment*. Generally, a deep learning model uses pooling layers to reduce the dimension of input data, this results in the heatmap calculated from the last Conv layer having a smaller size than the input data. To match the dimension between the learned heatmap and the input data, a linear mapping must be used for the alignment. For example, the VGG model is widely used for image processing [65]; it has a size of 224×224 for the input image, and the dimension of kernels in the last Conv layer is 14×14, which is one sixteenth of the size of the input image. Therefore, the VGG model needs to magnify the heatmap at sixteen times for each dimension, which indicates a large number of adjacent data points sharing the same value of heatmap, and thus reduces the resolution for the interpretation; (*ii*) *Weight sharing*. Other than the dimension alignment of the heatmap, weight sharing across different ECG leads in a deep learning model also affects the interpretive ability. For instance, the Conv kernels of DNN models in previous research learn kernel weights across all ECG leads [38], and it is therefore difficult to interpret each lead precisely using the shared weights. With these considerations, we develop the following techniques to obtain a refined resolution for the interpretation. In particular, we develop an *isolation-integration* strategy to allow the deep learning model to learn lead-wise weights from the ECG recording. This strategy is defined in the following steps:

i. At the isolation stage, in order to reduce the effect of weight sharing on the interpretation, we separate each of the 12 leads in an ECG recording, and use each isolated lead as an independent input to the model. This strategy allows the DNN model to learn features precisely for each separated ECG lead, rather than shared weights across multiple channels;
ii. At the integration stage, we develop a stepwise strategy to combine the learned features from each ECG lead, which enables the DNN model to explore elaborate relationships between different ECG leads, and prompts a comprehensive decision for diagnosis using the combined information. Some previous research used a global pooling layer for feature integration, however, the temporal information of the learned features would lost due to this global pooling [48];
iii. For the dimension alignment, it is important to ensure a similar size between the calculated heatmap and the input data, which will reduce the effect of data alignment on visualising salient features. As the dimension reduction of a DNN model is mostly from the pooling process, we therefore use a minimum number of pooling layers in the feature learning stage. This results in the kernel size of the last Conv layer having a close dimension to the input data, and therefore produces a refined resolution for the interpretation.

Using the above principles and utilising the residual deep neural networks [26, 38], we design a channel-wise residual deep learning model (CResNet). We present the details of developing the CResNet model in the next section.

#### 4.2.2 The CResNet Model and Network Training

Following our developed *isolation-integration* strategy, we first separate each of the 12 leads in the ECG recording, and use the isolated leads as inputs to the deep learning model; Then, we develop modules using residual neural networks (ResNet) to learn latent features for each ECG lead, as the ResNet has shown promising performance on processing ECG recordings in literature [26, 38]; Next, we use the long short-term memory model (LSTM) to learn temporal information, as well as the relationships between different ECG leads; Finally, the integrated features of the 12 ECG leads are used for the model prediction.

As illustrated in Extended Figure S2, our developed CResNet model has 12 channels for the model inputs, with each channel corresponding to one ECG lead. For each isolated input channel, we first use a Conv layer with 16 kernels to learn latent features from raw data of the ECG lead, which is followed by a batch normalisation (BN) layer, a rectified linear unit (ReLU) activation layer, and a max pooling layer. Next, we use four residual blocks to learn deep features from each lead, and each of the residual blocks consists of four repeated modules with the BN, ReLU, and Conv layers. In the first two residual blocks, the Conv layer has 16 kernels with a width size of 16; In the remaining two residual blocks, the Conv layer has 48 kernels and the width size of 48. After the second residual block, we use a Conv layer with 48 kernels to align dimensions with the following third residual block. At the end of each channel, we use a Conv layer with 48 kernels to finish feature learning for the ECG lead.

After learning features from the isolated input channels, the features are processed in the integration stage as illustrated in Extended Figure S2. We stepwise integrate the features to learn elaborate relationships between different ECG leads. We generate a feature matrix by concatenating the learned features from each of the isolated channels. As there is only one pooling layer used for each input channel in the isolation stage, the temporal dimension of the generated feature matrix in the concatenate layer is half the size of the input ECG recording. We note that the last Conv layer in each channel has the size of 48 kernels, and the generated feature matrix has a dimension of 576, which is obtained by concatenating Conv layers in the 12 ECG leads. Next, we learn relationships between different ECG leads using a bidirectional long short-term memory (BiLSTM) block and two time-distributed dense layer (TD Dense) blocks. Both the BiLSTM block and TD Dense blocks consist of a max pooling layer (MaxP), an average pooling layer (AvgP), and a dropout layer. The BiLSTM block consists of two LSTM layers, one has a forward direction and the other has a reverse direction, and each LSTM layer has 64 cells in the hidden state. For the two TD Dense blocks, the first one has 64 units and the second block has 32 units. We then flatten layers of the TD Dense block followed by a fully connected layer with 128 units. Finally, we use a sigmoid function to calculate probability for the output of model prediction.

The developed CResNet model is used to perform the three tasks in this study, i.e., ECG abnormality diagnosis, gender identification, and hypertension screening. We train the CResNet model independently for each of the three tasks, whilst keep the model architecture and hyperparameters the same for all the three tasks, i.e., the number of neurons, activation function, optimizer, batch size, and epochs. For the first task, the CResNet model has an output vector of six values, indicating the six types of ECG abnormalities; For the second task, the CResNet model has an output of a single value, indicating the probability of male or female; For the third task, the CResNet model also has an output of a single value, indicating the probability of hypertension presented for the subject.

The neural network was trained using the loss of binary cross-entropy, which was minimized by the Adam optimizer with default parameters [66]. Hyper-parameters of the network architecture were chosen via a combination of grid search and manual tuning with the following considerations, the number of residual blocks {2, 4, 8}, kernel size for the Conv layer {16, 32, 48, 64}, the number of BiLSTM blocks {1, 2, 4}, the size of pooling layers {2, 4}, dropout rate of {0, 0.2, 0.5, 0.6}, the mini batch size of {32, 64, 128}, initial learning rate of {10^−2^, 10^−3^, 10^−4^}, the number of epochs without improvement in plateaus between 7 and 15, which would result in a reduction of the learning rate by a factor of 10. After tuning the parameters with 300K samples a small scale of the dataset, we set a learning rate of 10^−4^ and use the whole dataset to train the model with a mini batch size of 128 samples, and the maximum number of epochs was set as 70. During the model training, a holdout set with 10% of the data was used for the validation. We tried different configurations of the model development, especially in the feature integration stage, such as the BiLSTM, LSTM, and TD Dense layers; and found that the combination of BiLSTM with two TD Dense layers shows good performance for the diagnosis. To reduce the effect of imbalanced classes in the dataset, we weighted each sample by multiplying a score of 2*log(*n*_ECGs_/*n*_Class_), where *n*_ECGs_ indicates the total number of samples, and *n*_Class_ is the size of samples in the class. A total of 20 Nvidia V100 GPUs in a high performance computing platform are available to train the CResNet model, which is located at the Institute of Biomedical Engineering, Department of Engineering Science, University of Oxford.

#### 4.2.3 The Interpretation Model and Visualisation

The interpretation model is paired with the CResNet model, and it is used in post-hoc analysis to visualise salient features that have been used by the CResNet model for decision making. Figure S3 shows the flowchart of developing the interpretation model. For each of the 12 leads in an ECG recording, the data is first processed using the ResNet model with our proposed isolation strategy, and a feature matrix can be obtained by concatenating features from all of the 12 ECG leads; Then, the concatenated feature matrix is processed in the integration stage and used for the model prediction; Next, we use back propagation to compute gradient for the prediction with respect to the concatenated feature matrix, and the gradient matrix can be obtained accordingly. Keep in mind, the concatenated feature matrix is computed using an isolated strategy, therefore the calculated gradient matrix can precisely weight Conv kernels for each of the 12 ECG leads. Next, we weight kernels in the concatenated feature matrix using the averaged gradient scores, which are then filtered by a ReLu function. Finally, the heatmap of feature importance can be obtained by performing the dimension alignment. Notably, because the concatenated feature matrix has a half size of the input ECG recording in the temporal direction, the calculated heatmap only needs to be magnified two times for the dimension alignment, which allows to produce salient features with a refined resolution for the interpretation of the ECG recording.

### 4.3 Statistical and Empirical Analysis of Model Performance

To evaluate the performance of our developed CResNet model on the three tasks, we calculate standard matrices of the testing results for each independent task. We compute the area under the receiver operating characteristic curve (AUC-ROC) to report the model performance; we also calculate the F1-score for the first task, as it has an imbalanced testing dataset, and the score is used to compare the performance of our model with the evaluation results from the cardiology professionals and the state-of-the-art model [38]. We calculate the micro average across different classes to report an overall score of the model performance, which computes the total true positives, false negatives, and false positives to obtain a comprehensive metric. The optimal cutoff point for the sensitivity and specificity scores is obtained by maximising the G-mean value, which is a geometric mean of the two scores [67]. We use the diagnostic odds ratio (DOR) to indicate the model’s ability of diagnosis, which is calculated as the positive likelihood ratio (sensitivity / (1-specificity)) to the negative likelihood ratio ((1-sensitivity) / specificity). A value of DOR larger than 1 indicates the model having the discriminatory test performance, with the DOR value correlating positively with better diagnosis performance [68]. We use the bootstrap method (repeated sampling for 1,000 times) to compute the 95% confidence interval (CI) and standard deviation for the calculated indices [21, 38]. We use two-sided McNemar’s *χ*^2^ test to evaluate differences between classification results for paired samples [38, 69, 70], and use Pearson’s *χ*^2^ test to evaluate differences for unpaired samples [71]. We also calculate Cohen’s kappa coefficient to test inter-rater/-model agreement [72]. We consider a *p*-value of less than 0.05 as statistically significant.

## Acknowledgment

The research was partially supported by the National Institute for Health Research (NIHR) Oxford Biomedical Research Centre (BRC). The views expressed are those of the author(s) and not necessarily those of the NHS, the NIHR or the Department of Health. T.Z. was supported by the RAEng Engineering for Development Research Fellowship. A.L.P.R. was supported in part by CNPq (310790/2021-2 and 465518/2014-1) and by FAPEMIG (PPM-00428-17 and RED-00081-16); A.H.R. was partially supported by the Kjell och Märta Beijer Foundation.

## Data Availability

As the original dataset of ECG recordings is prohibitively large for upload, about 15% of the CODE dataset has been made openly available as annotations (https://doi.org/10.5281/zenodo.4916206), including 345,779 ECG recordings collected from 233,770 subjects. The standard testing dataset and the labels are also publicly accessible (https://zenodo.org/record/3765780#.YbIaypHP36c).

## Author Contributions

L.L., T.Z., E.Z., and D.A.C. contributed to design the study; A.H.R. and A.L.P.R. collected the experimental data; L.L. and T.Z. ran the experiments; L.L. created the figures and tables; L.L., T.Z., A.H.R., and E.Z. contributed to data analysis; L.L., T.Z., A.H.R., L.C., E.Z., A.L.P.R., Y.Z., and D.A.C. contributed to the discussion; D.A.C., A.L.P.R., and Y.Z. were senior advisors of the project. All authors read the manuscript and approved the submission.

## Extended Data Figures and Tables

**Extended Figure S1:**
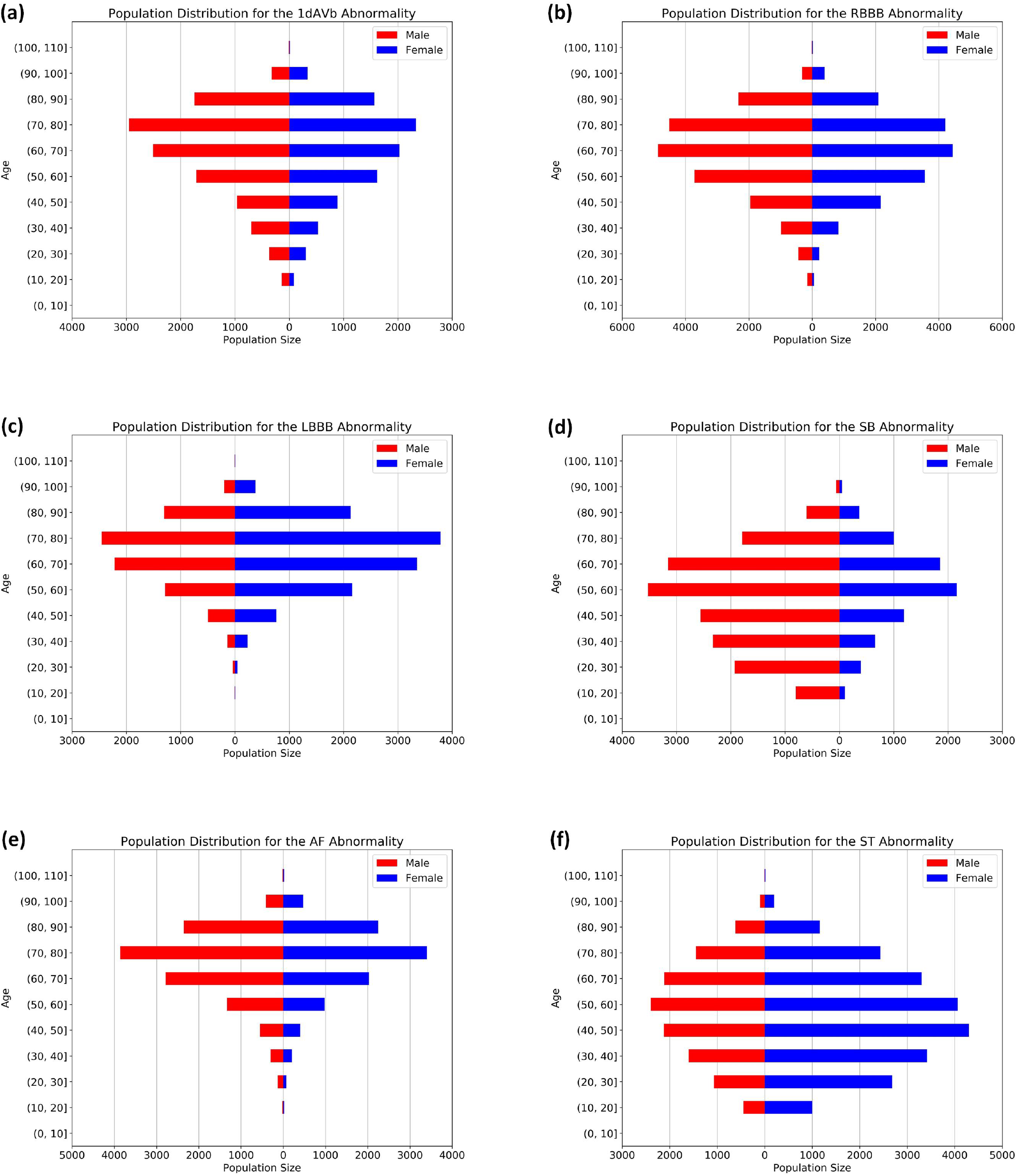
Prevalence of ECG abnormalities in the CODE dataset (*n*_Subjects_ = 1,558,772), including **(a)** first-degree atrioventricular block (1dAVb), **(b)** right bundle branch block (RBBB), **(c)** left bundle branch block (LBBB), **(d)** sinus bradycardia (SB), **(e)** atrial fibrillation (AF), and **(f)** sinus tachycardia (ST).

**Extended Table S1:**
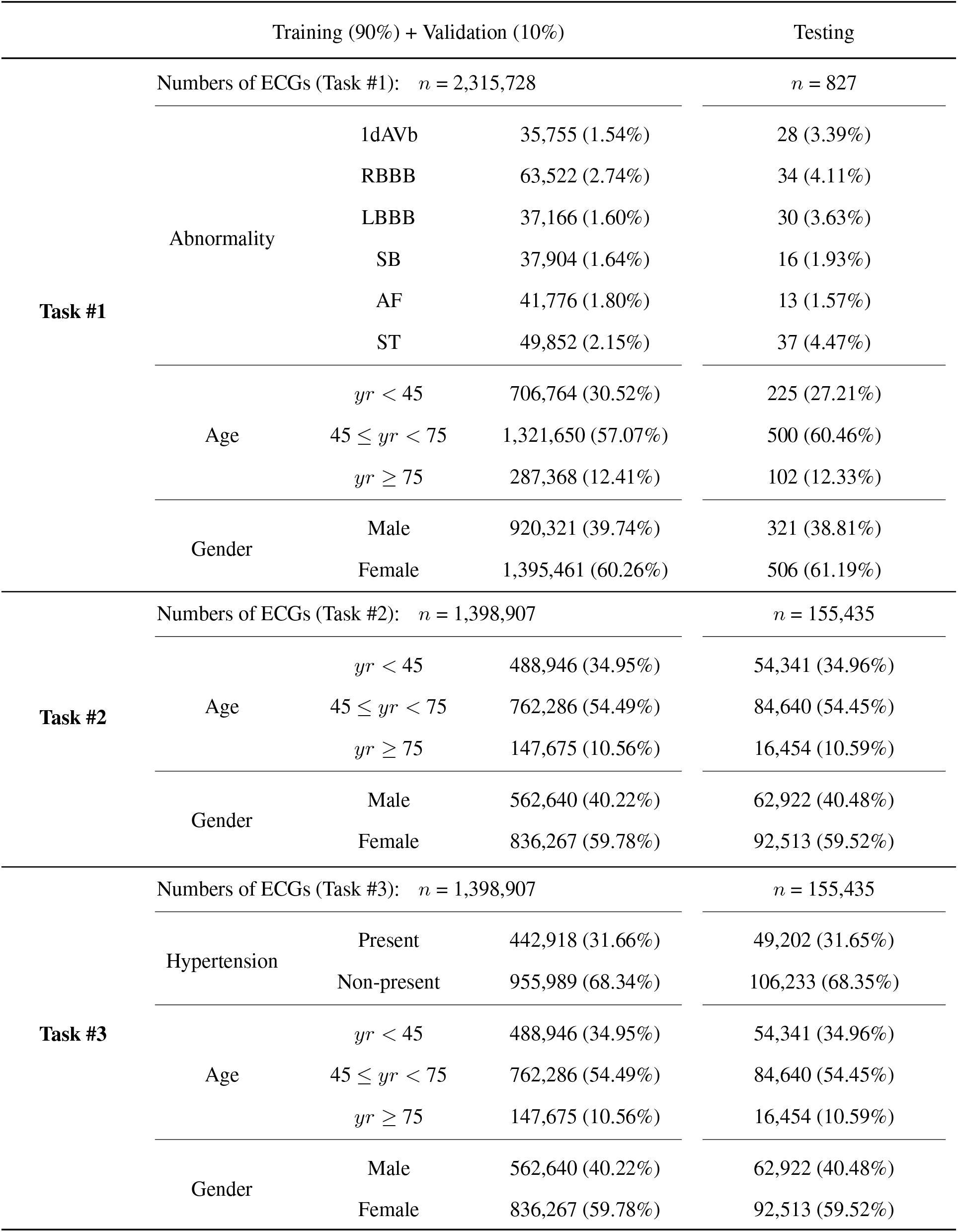
Dataset and study population for the three tasks in this study (Numbers and percentages).

**Extended Figure S2:**
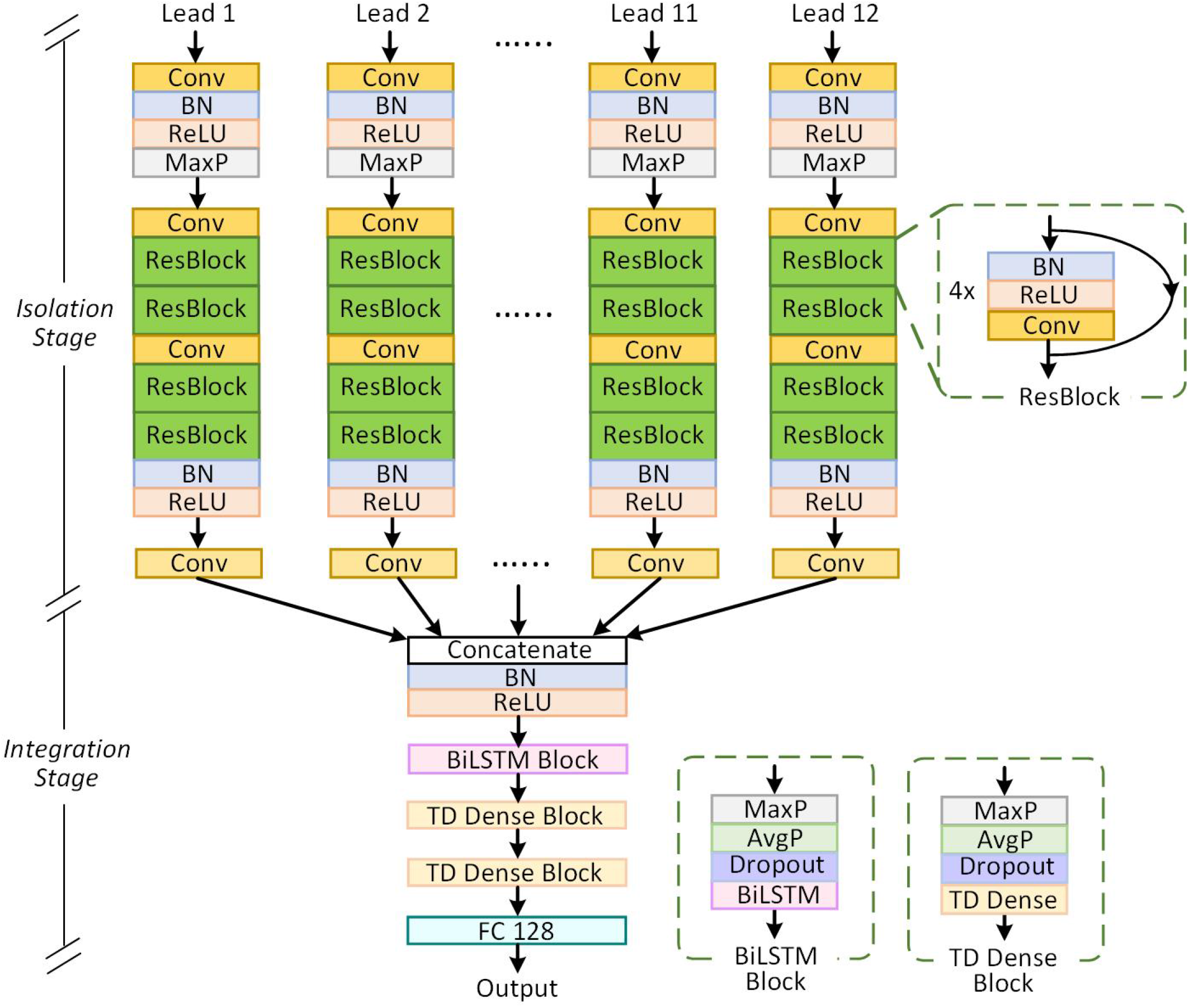
The developed channel-wise residual deep neural networks (CResNet) for this study.

**Extended Figure S3:**
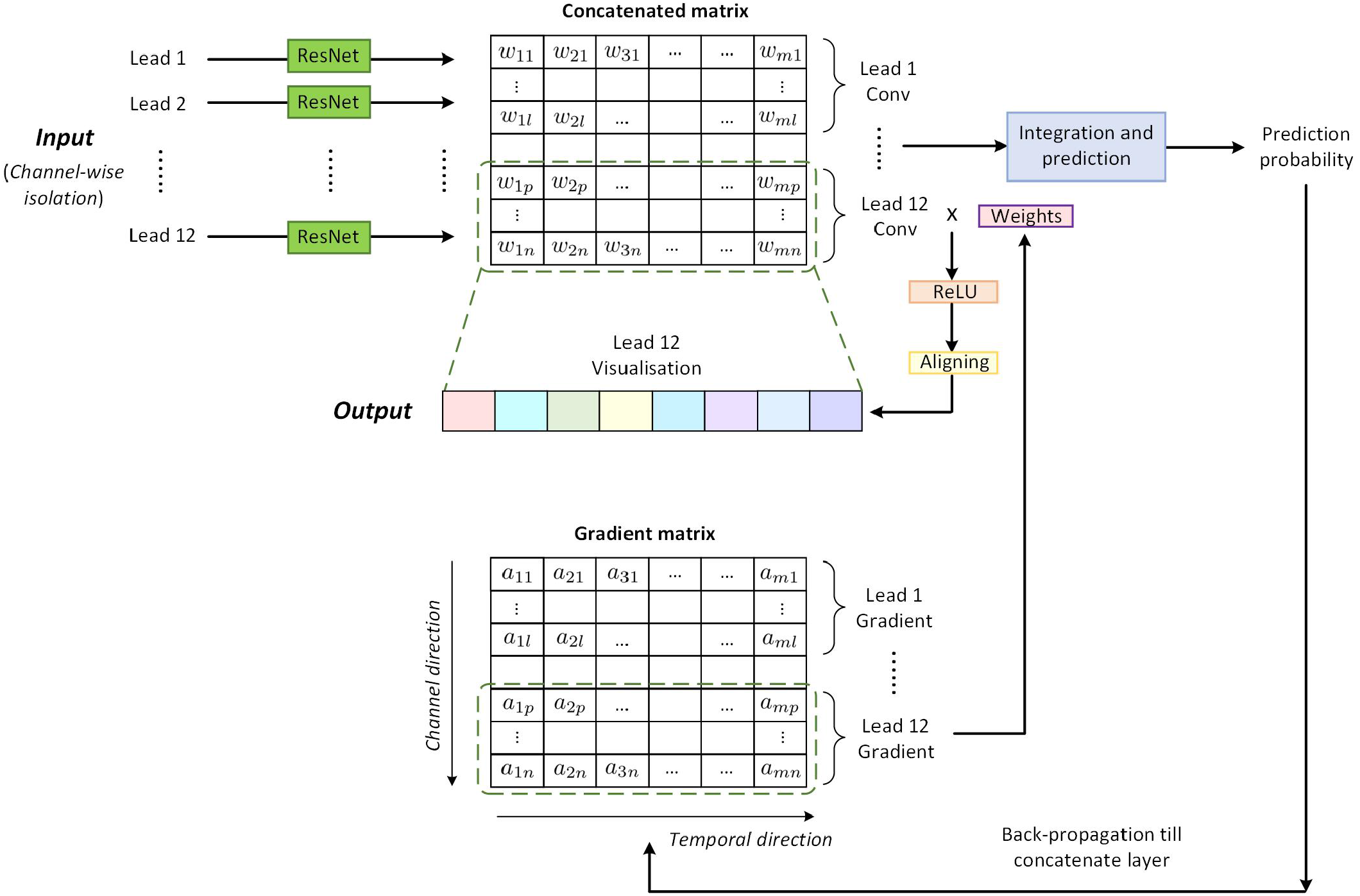
The developed interpretation module for visualising salient features using Grad-CAM method. The figure illustrates the pipeline of visualising salience information in the 12^*th*^ lead of an ECG recording.

**Extended Figure S4:**
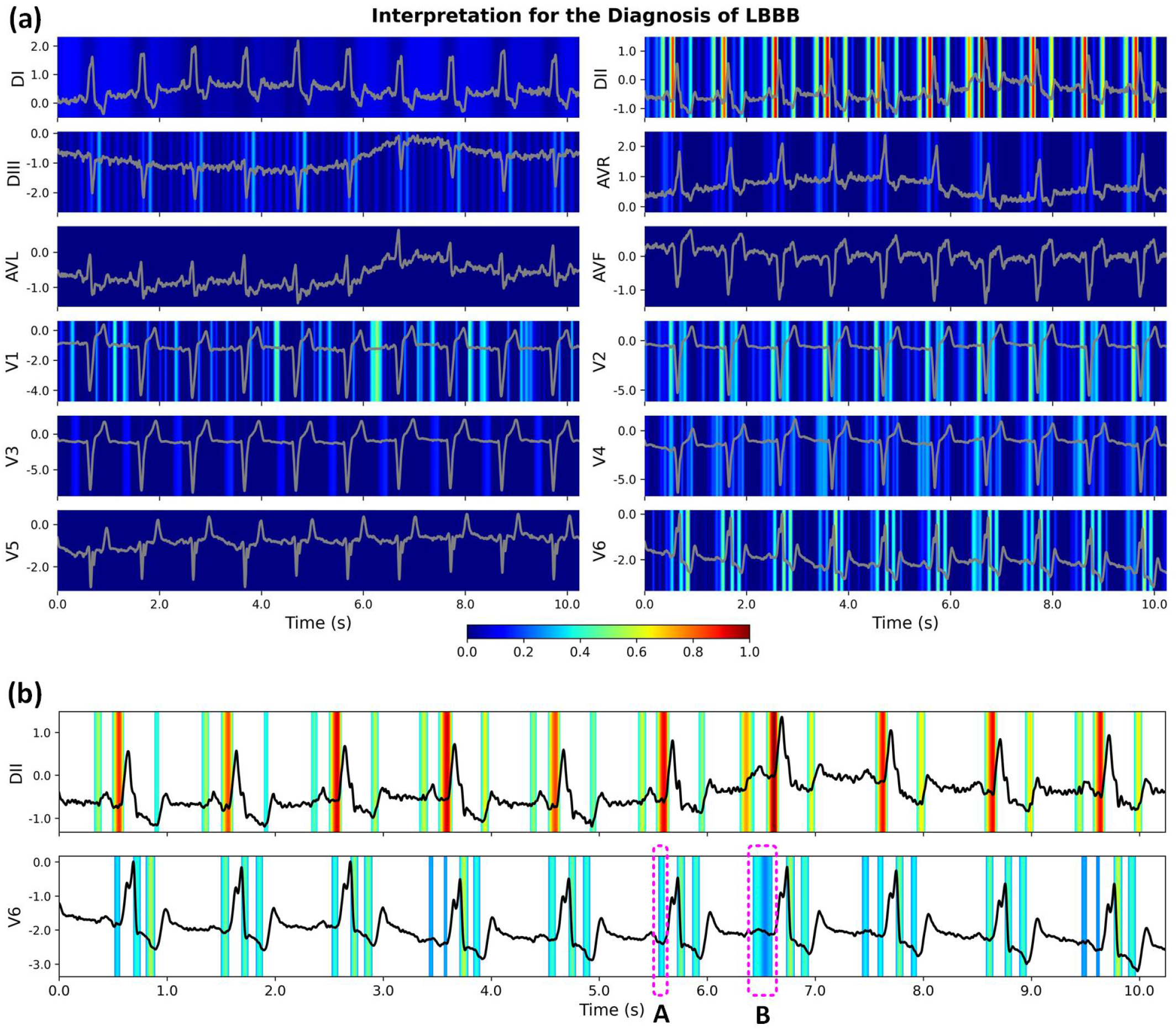
Interpretation for the diagnosis of left bundle branch block (LBBB) using the proposed CResNet model. **(a)** The original calculated heatmaps for 12 ECG leads, with colour bar ranging from blue to red indicating the increasing weights of importance. **(b)** The refined view of heatmaps for the DII and V6 leads with background colour removed. The diagnosis criteria of LBBB include the absence of Q waves in lateral leads, notched R waves in lateral leads, and T wave inversion [50, 51]. Our proposed CResNet model recognises these pathological morphologies successfully in the V6 lead. The model also identifies other salient waves in the DII lead, such as the absence of Q waves, T inversion, and the segments before P waves. Using the combination of salient waves in the 12 ECG leads, the CResNet model diagnoses the LBBB with the probability of 0.974; While with the removal of the DII lead, we obtained a prediction probability of 0.886, indicating additional knowledge derived from DII was missing. Notably, our model is very flexible in identifying the pathological morphologies. For example, the morphologies in segments *A* and *B* have different time durations, and the CResNet model identifies the varying lengths for the two segments successfully in the V6 lead.

**Extended Figure S5:**
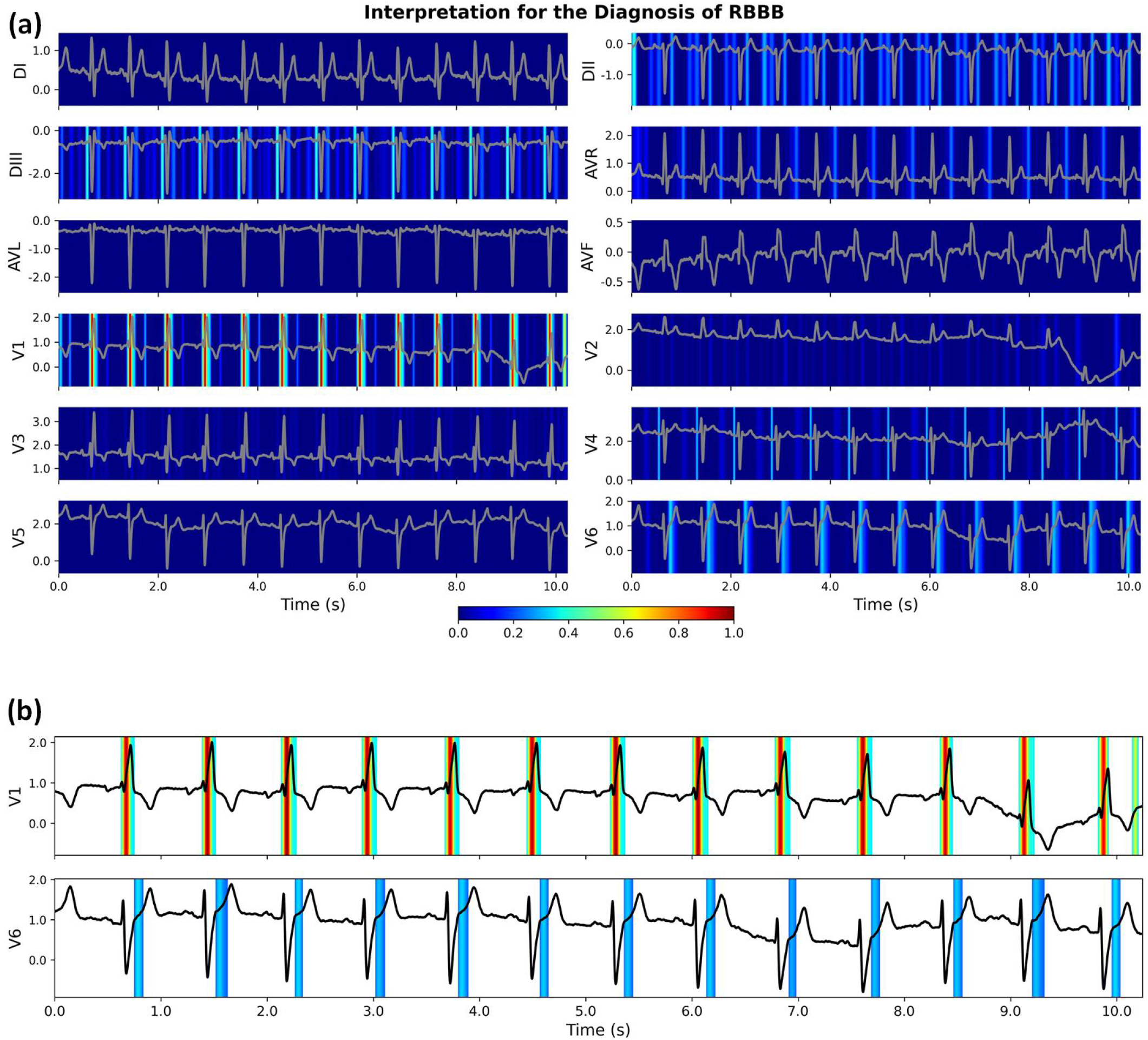
Interpretation for the diagnosis of right bundle branch block (RBBB) using the proposed CResNet model.**(a)** The original calculated heatmaps for 12 ECG leads, with colour bar ranging from blue to red indicating the increasing weights of importance. **(b)** The refined view of heatmaps for V1 and V6 leads with background colour removed. For the diagnosis of RBBB, the RSR^*′*^ pattern in the anterior precordial leads is an important criterion [50, 51]. Our proposed CResNet model recognises the ‘M-shaped’ QRS complexes successfully in the V1 lead, and it also highlights the importance of J waves in the V6 lead. Using the combined salient features, the CResNet model has a probability of 0.929 to diagnose the RBBB with the ECG recording.

**Extended Figure S6:**
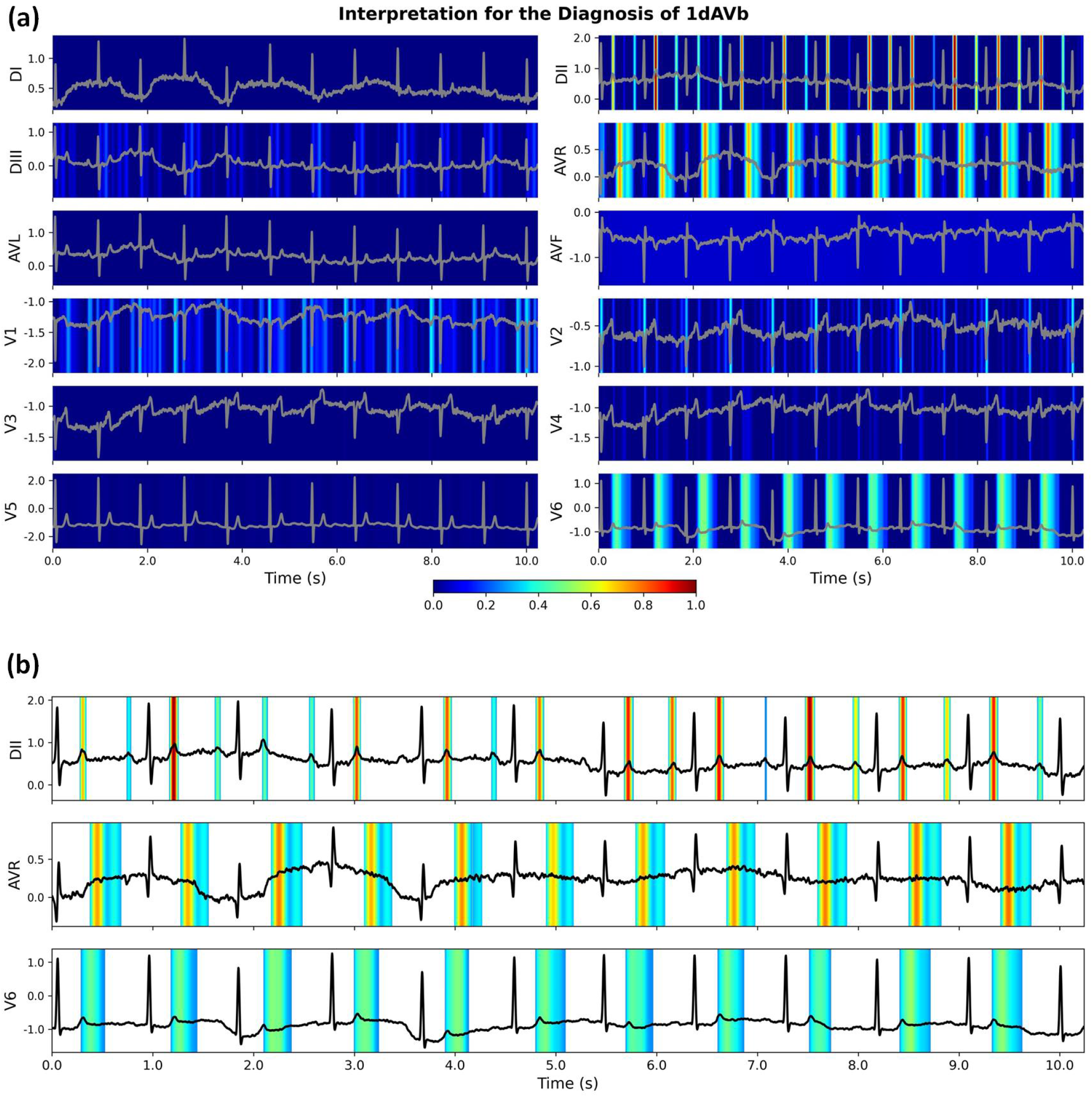
Interpretation for the diagnosis of first degree atrioventricular block (1dAVb) using the proposed CResNet model. **(a)** The original calculated heatmaps for 12 ECG leads, with colour bar ranging from blue to red indicating the increasing weights of importance. **(b)** The refined view of heatmaps for DII, AVR, and V6 leads with background colour removed. For the diagnosis of 1dAVb, the criteria include prolonged PR interval, normal QRS, and normal rhythm [50, 51]. Because the QRS complex is normal, our proposed CResNet model highlights other morphologies for the diagnosis, such as the T and P waves in the DII lead, and the segments after T waves in the AVR and V6 leads. Using the combination of salient features, the CResNet model has a probability of 0.932 to diagnose the 1dAVb with the ECG recording.

**Extended Figure S7:**
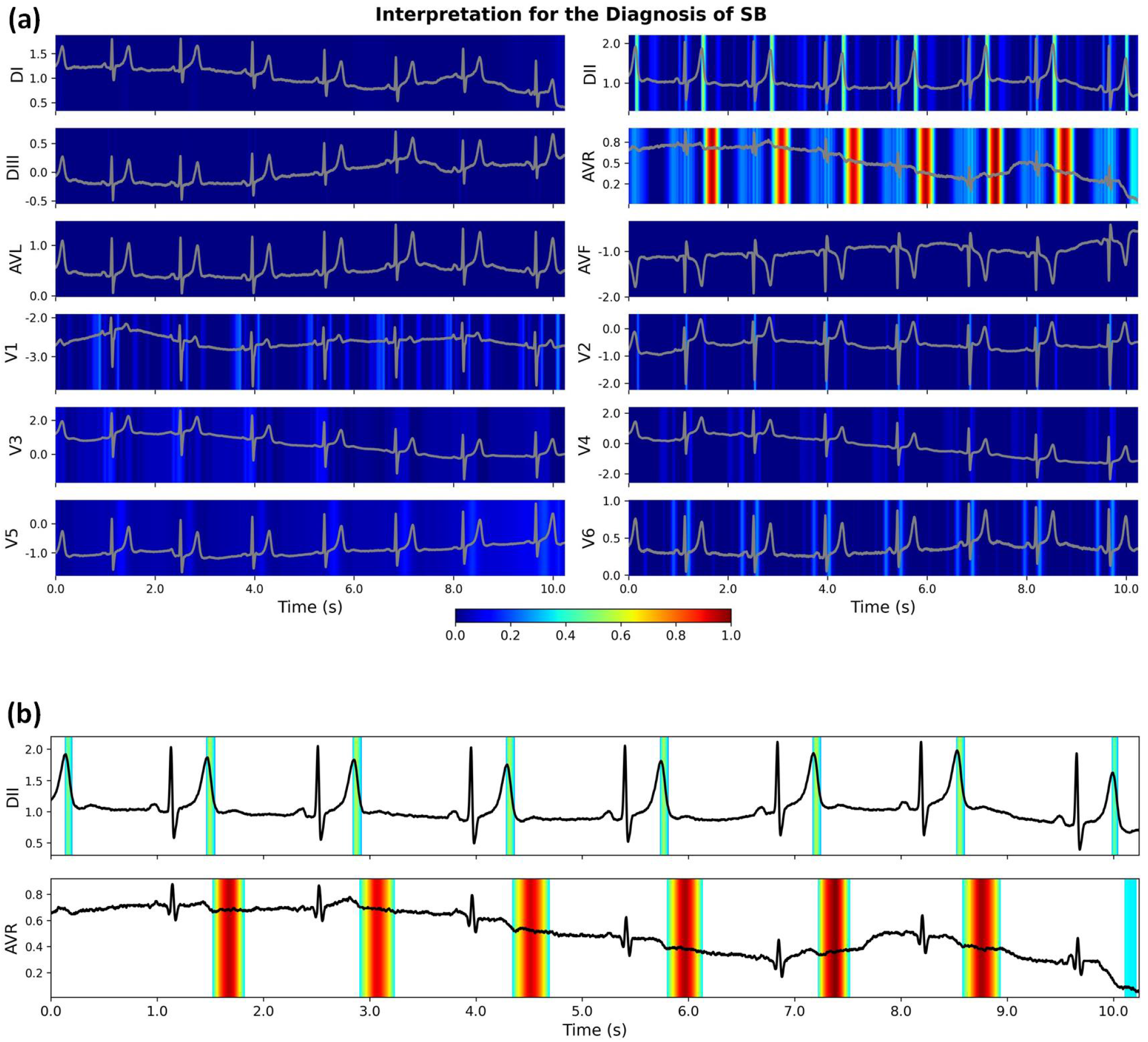
Interpretation for the diagnosis of sinus bradycardia (SB) using the proposed CResNet model. **(a)** The original calculated heatmaps for 12 ECG leads, with colour bar ranging from blue to red indicating the increasing weights of importance. **(b)** The refined view of heatmaps for the DII and AVR leads with background colour removed. SB is defined as a sinus rate below 50 bpm with otherwise normal P, QRS and T waves [50, 51]; while the prominent U waves were also reported for asymptomatic SB in literature [42]. For the diagnosis of SB, our proposed CResNet model highlights the U waves in the AVR lead and the downslops of T waves in the DII lead. Using the combination of salient features, the CResNet model has a probability of 0.932 to diagnose the SB using the ECG recording.

**Extended Figure S8:**
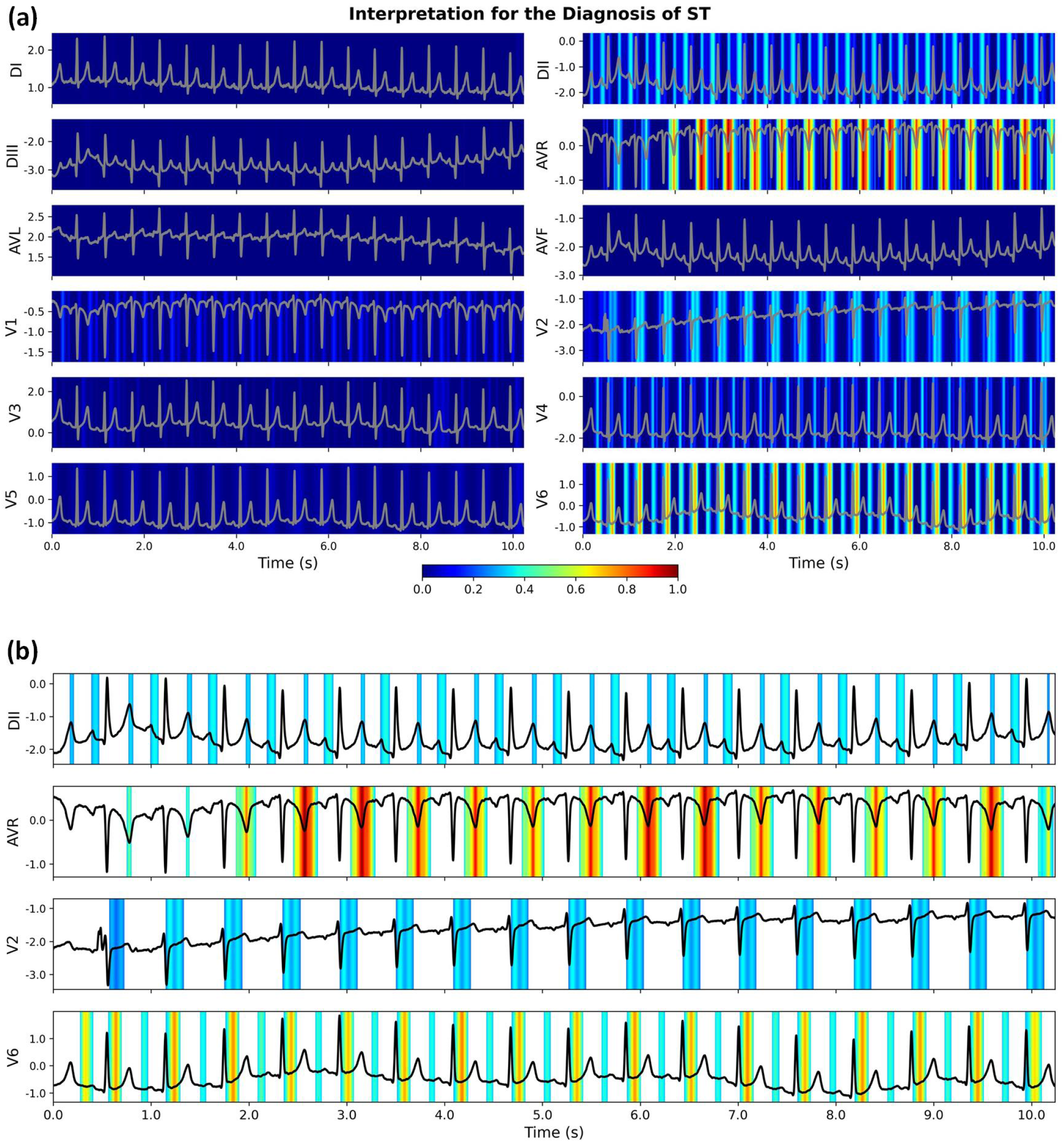
Interpretation for the diagnosis of sinus tachycardia (ST) using the proposed CResNet model. **(a)** The original calculated heatmaps for 12 ECG leads, with colour bar ranging from blue to red indicating the increasing weights of importance. **(b)** The refined view of heatmaps for the DII, AVR, V2, and V6 leads with background colour removed. ST is the sinus rhythm with a heart rate greater than 100/min, and it has normal P wave preceding every QRS complex [50, 51]. For the diagnosis of ST, our proposed CResNet model highlights the downslops of T and P waves in the DII lead, T waves in the AVR lead, and ST segments in the V2 and V6 leads. Using the combination of salient features, the CResNet model has a probability of 0.948 to diagnose the ST with the ECG recording.

**Extended Figure S9:**
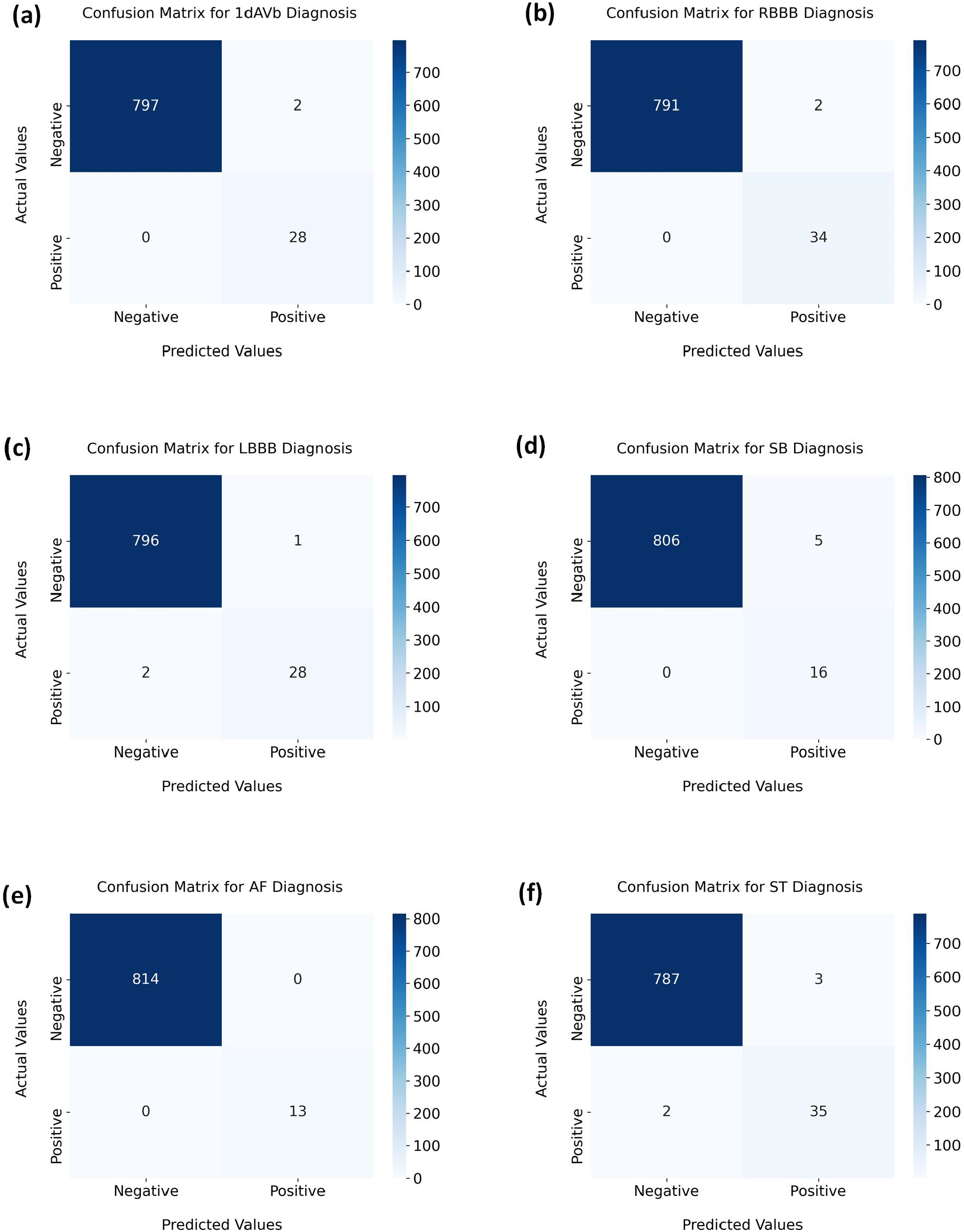
Confusion matrices for the diagnosis of ECG abnormalities in the holdout testing dataset using 12-lead ECGs, including **(a)** first-degree atrioventricular block (1dAVb), **(b)** right bundle branch block (RBBB), **(c)** left bundle branch block (LBBB), **(d)** sinus bradycardia (SB), **(e)** atrial fibrillation (AF), and **(f)** sinus tachycardia (ST).

**Extended Table S2:** Performance comparison for the diagnosis of ECG abnormalities with the McNemar’s test.

| | | McNemar’s $\chi^2$ test ($p$-value) | | | | | |
| --- | --- | --- | --- | --- | --- | --- | --- |
|  |  | 1dAVb | RBBB | LBBB | SB | AF | ST |
| CResNet | vs Cardio. Rd. | 4.9231<br>**(0.0265)** | 1.5000<br>(0.2207) | 0.2500<br>(0.6171) | 0.0000<br>(1.0000) | 4.1667<br>**(0.0412)** | 0.4444<br>(0.5050) |
| CResNet | vs Emerg. Rd. | 12.5000<br>**(0.0004)** | 3.2727<br>(0.0704) | 0.1667<br>(0.6831) | 0.1250<br>(0.7237) | 5.1429<br>**(0.0233)** | 0.0000<br>(1.0000) |
| CResNet | vs Medical Sd. | 12.1905<br>**(0.0005)** | 0.8000<br>(0.3711) | 0.1250<br>(0.7237) | 0.4444<br>(0.5050) | 8.1000<br>**(0.0044)** | 0.9000<br>(0.3428) |
| CResNet | vs Cardio. #1 | 4.0833<br>**(0.0433)** | 0.0000<br>(1.0000) | 0.0000<br>(1.0000) | 0.1250<br>(0.7237) | 1.3333<br>(0.2482) | 0.0833<br>(0.7728) |
| CResNet | vs Cardio. #2 | 0.1667<br>(0.6831) | 0.5000<br>(0.4795) | 1.3333<br>(0.2482) | 0.1250<br>(0.7237) | 1.3333<br>(0.2482) | 0.1000<br>(0.7518) |
| CResNet | vs DNN_Comp [38] | 1.1250<br>(0.2888) | 0.5000<br>(0.4795) | 1.3333<br>(0.2482) | 0.0000<br>(1.0000) | 1.3333<br>(0.2482) | 0.5000<br>(0.4795) |

\*The table shows the two-sided McNemar’s $\chi^2$ test [38, 69, 70] and $p$ -values for the performance comparison between our proposed CResNet model and other evaluation results, including Cardio. Rd.: 4<sup>th</sup> year cardiology residents; Emerg. Rd.: 3<sup>rd</sup> year emergency residents; Medical Sd.: 5<sup>th</sup> year medical students; Cardio. #1: the 1<sup>st</sup> cardiologist; Cardio. #2: the 2<sup>nd</sup> cardiologist; DNN\_Comp: the state-of-the-art benchmark model developed in [38]. The bold-faced values denote statistical significance ( $p < 0.05$ ) for the comparison of paired evaluation results.

**Extended Table S3:**
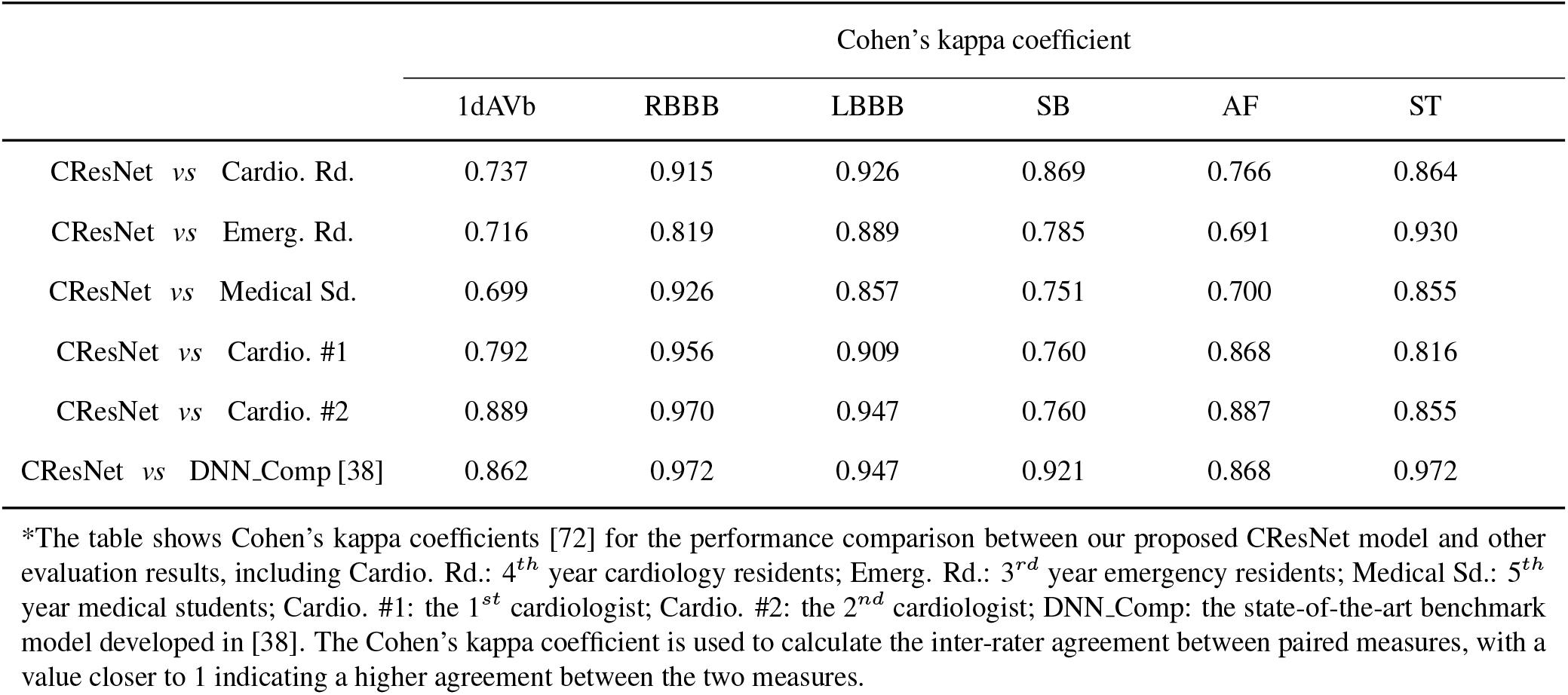
Performance comparison for the diagnosis of ECG abnormalities with Cohen’s kappa coefficient.

**Extended Table S4:**
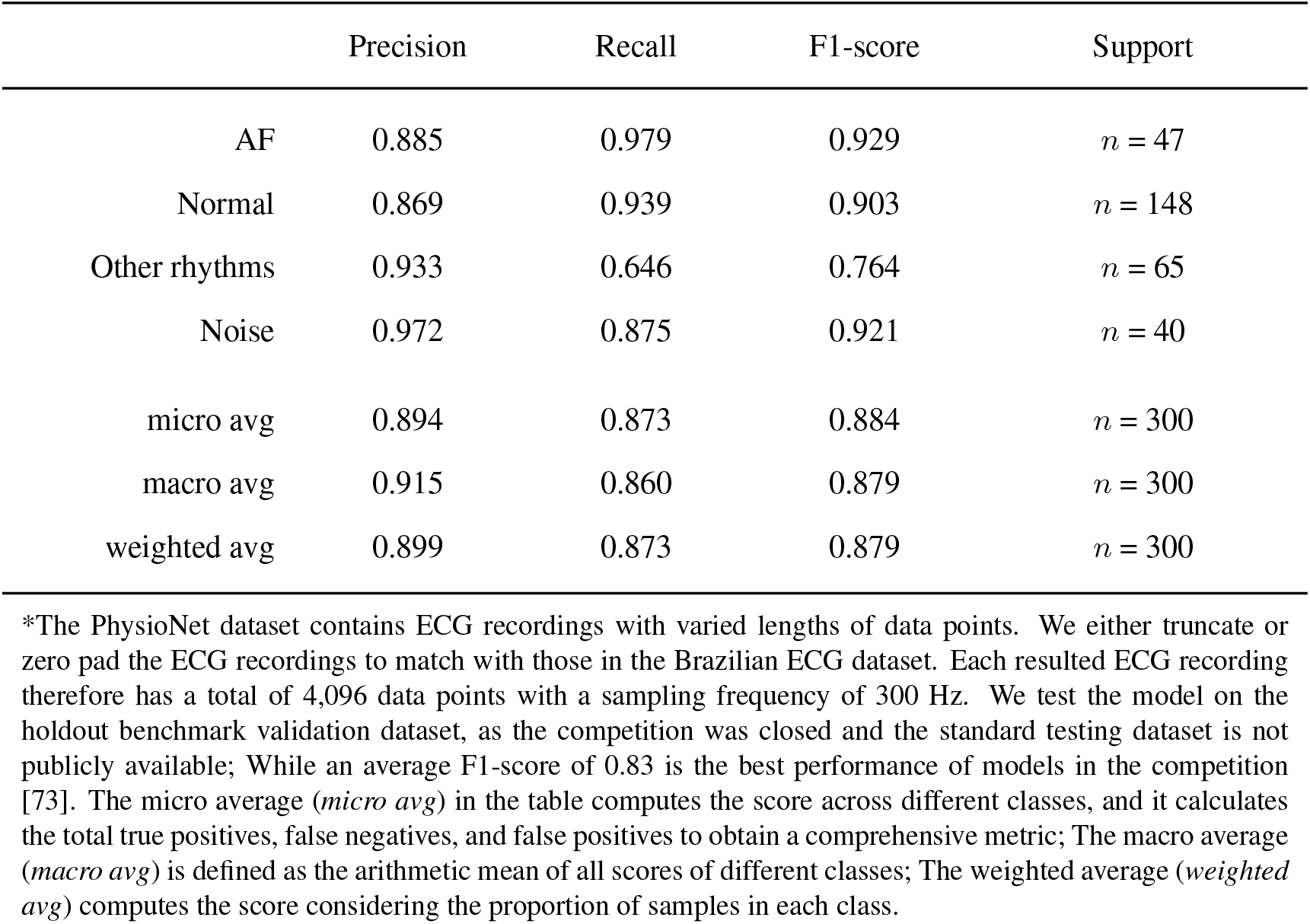
Model performance on the external dataset retrieved from the PhysioNet/CinC 2017 Challenge.

|  | Precision | Recall | F1-score | Support |
| --- | --- | --- | --- | --- |
| AF | 0.885 | 0.979 | 0.929 | $n = 47$ |
| Normal | 0.869 | 0.939 | 0.903 | $n = 148$ |
| Other rhythms | 0.933 | 0.646 | 0.764 | $n = 65$ |
| Noise | 0.972 | 0.875 | 0.921 | $n = 40$ |
| micro avg | 0.894 | 0.873 | 0.884 | $n = 300$ |
| macro avg | 0.915 | 0.860 | 0.879 | $n = 300$ |
| weighted avg | 0.899 | 0.873 | 0.879 | $n = 300$ |

\*The PhysioNet dataset contains ECG recordings with varied lengths of data points. We either truncate or zero pad the ECG recordings to match with those in the Brazilian ECG dataset. Each resulted ECG recording therefore has a total of 4,096 data points with a sampling frequency of 300 Hz. We test the model on the holdout benchmark validation dataset, as the competition was closed and the standard testing dataset is not publicly available; While an average F1-score of 0.83 is the best performance of models in the competition [73]. The micro average (*micro avg*) in the table computes the score across different classes, and it calculates the total true positives, false negatives, and false positives to obtain a comprehensive metric; The macro average (*macro avg*) is defined as the arithmetic mean of all scores of different classes; The weighted average (*weighted avg*) computes the score considering the proportion of samples in each class.

**Extended Figure S10:**
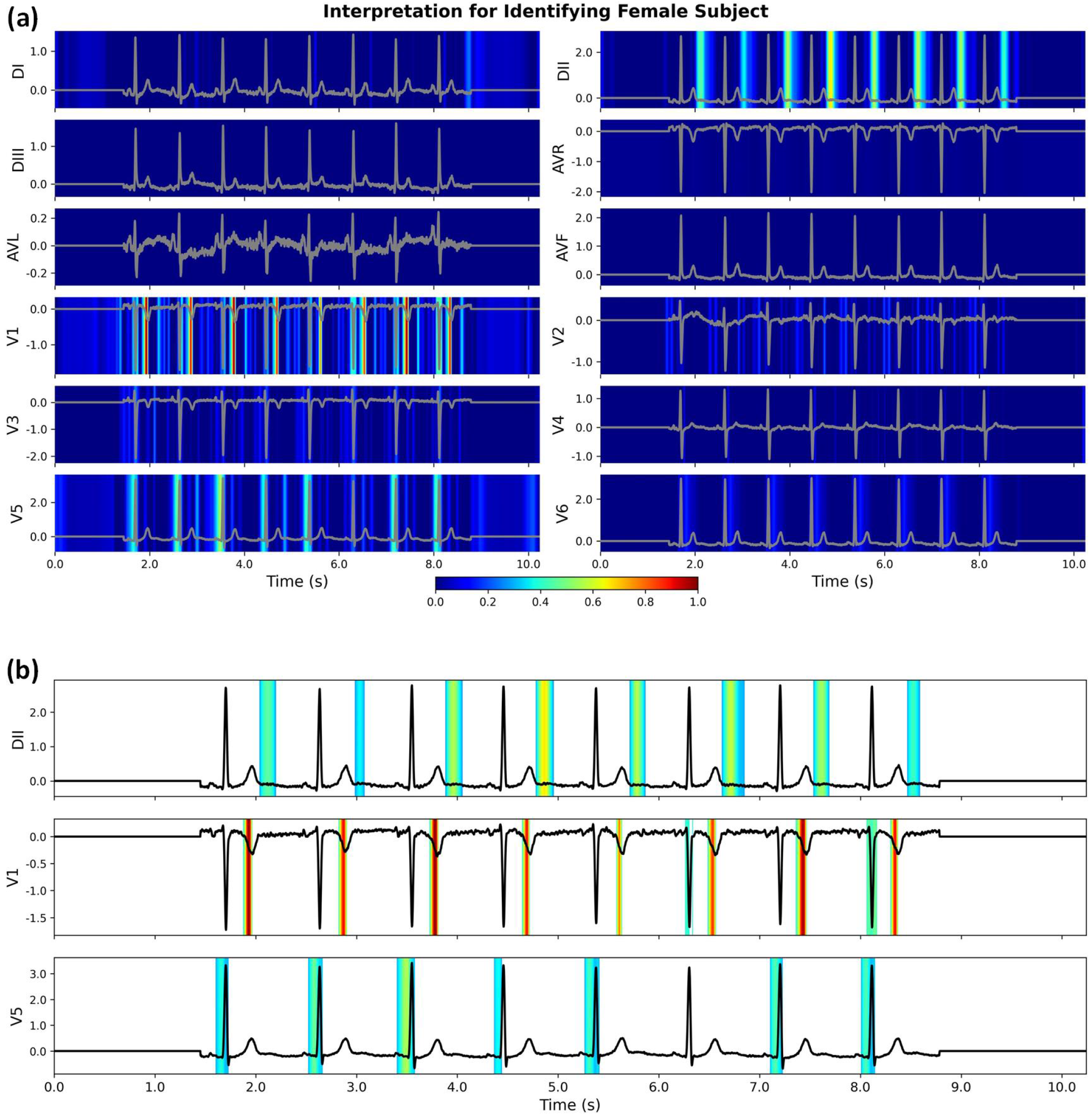
Interpretation for identifying female subject using our developed CResNet model. **(a)** The original calculated heatmaps for 12 ECG leads, with colour bar ranging from blue to red indicating the increasing weights of importance.**(b)** The refined view of the DII, V1, and V5 leads with background colour removed. Using the combination of salient features, the CResNet model has a probability of 0.971 to identify gender for the female subject using the ECG recording.

**Extended Figure S11:**
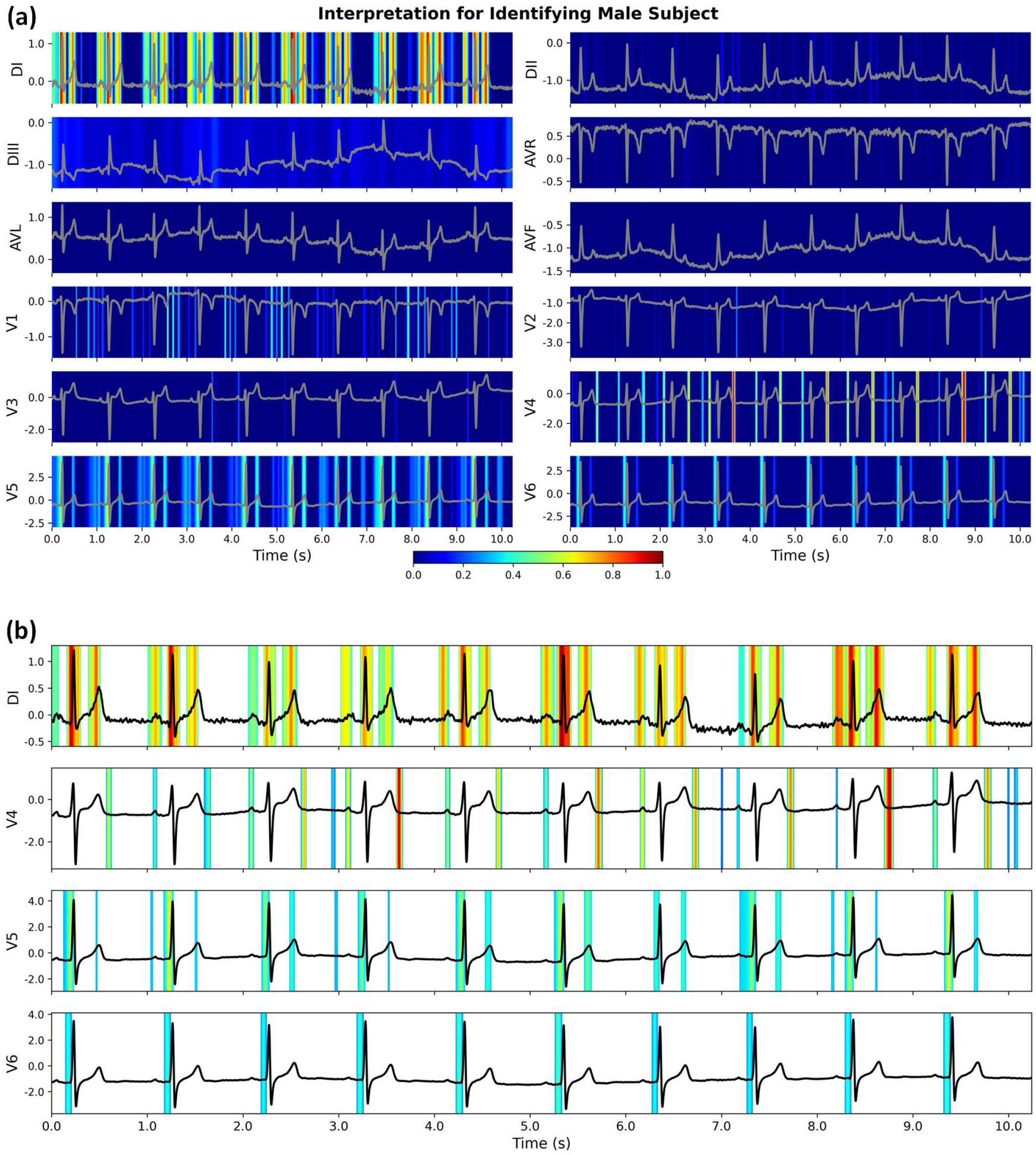
Interpretation for identifying male subject using our developed CResNet model. **(a)** The original calculated heatmaps for 12 ECG leads, with colour bar ranging from blue to red indicating the increasing weights of importance. **(b)** The refined view of the DI, V4, V5, and V6 leads with background colour removed. Using the combination of salient features, the CResNet model has a probability of 0.981 to identify gender for the male subject using the ECG recording.

**Extended Figure S12:**
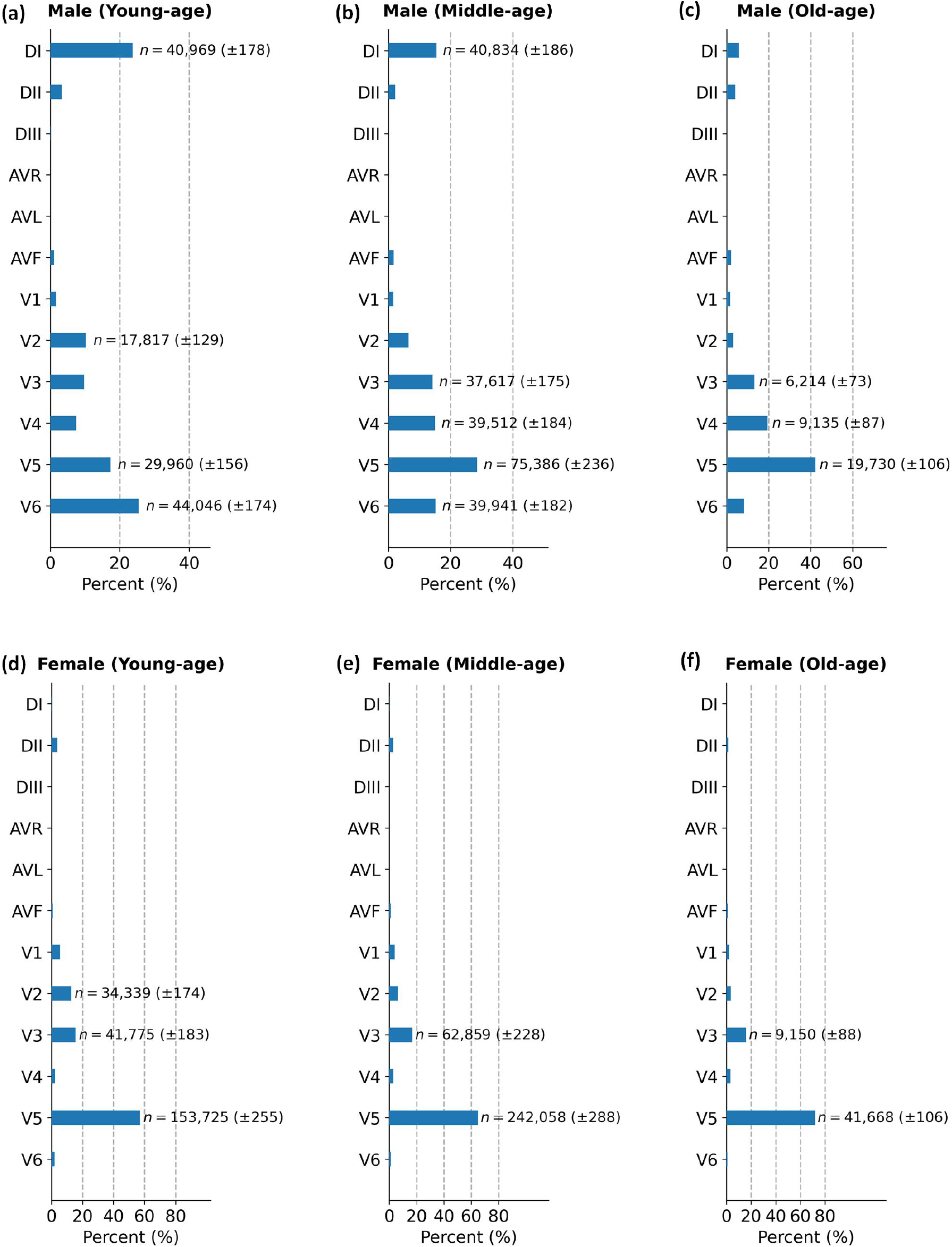
Distributions of dominant leads for gender identification using our developed CResNet model. **(a)** Distribution for young-age male subjects (*yr <* 45). **(b)** Distribution for middle-age male subjects (45 ≤ *yr <* 75). **(c)** Distribution for old-age male subjects (*yr* ≥ 75). **(d)** Distribution for young-age female subjects (*yr <* 45). **(e)** Distribution for middle-age female subjects (45 ≤ *yr <* 75). **(f)** Distribution for old-age female subjects (*yr* ≥ 75). We annotate the number of occurrences when the dominant lead accounts for more than 10% of all the 12 ECG leads. The number of occurrences is presented as mean and standard deviation.

**Extended Figure S13:**
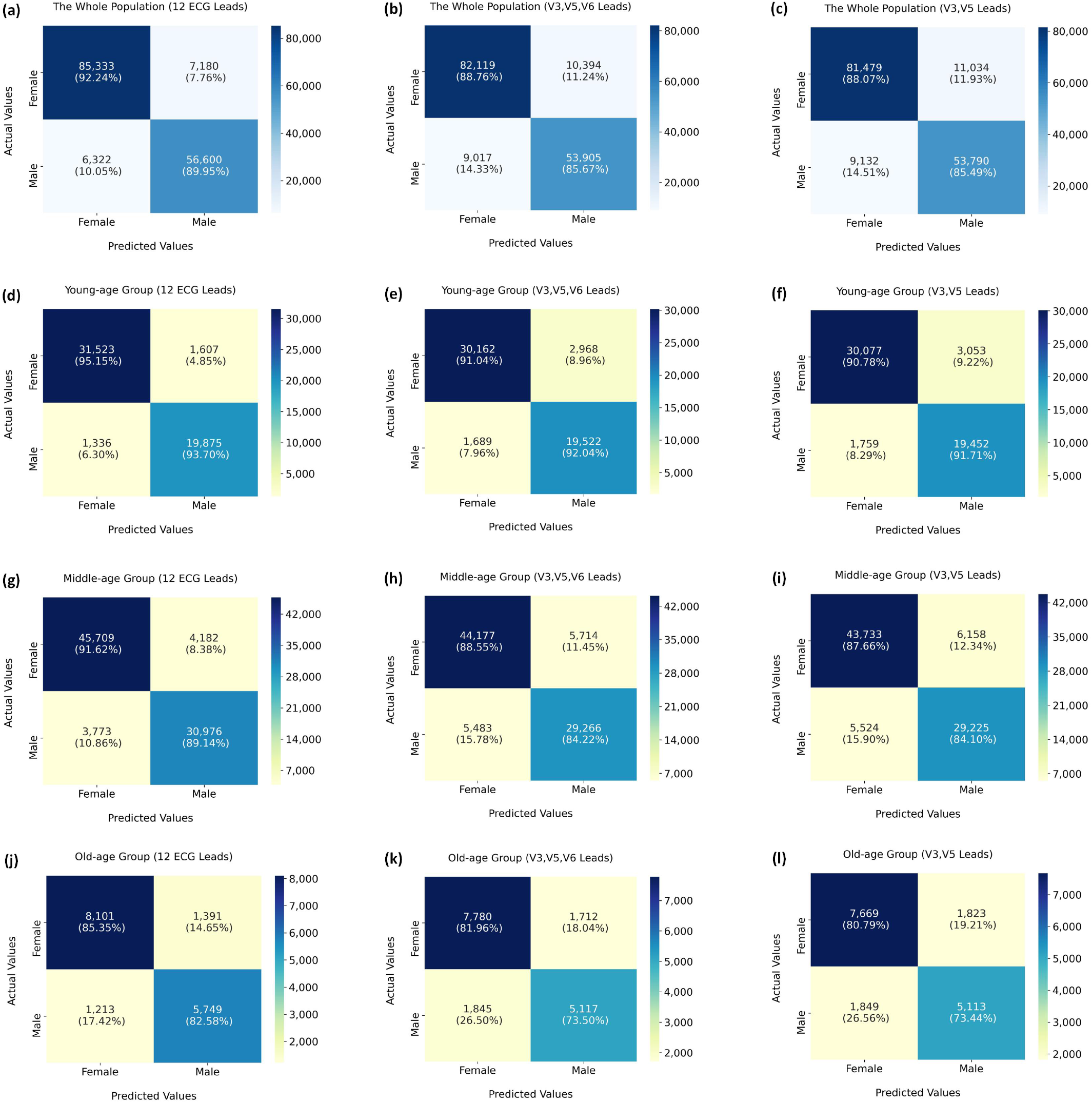
Confusion matrices of gender identification for the CResNet model using 12 ECG leads, dominant V3, V5, and V6 leads, and dominant V3 and V5 leads. **(a)**-**(c)** Performance comparison in the whole population. **(d)**-**(f)** Performance comparison young-age group (*yr <* 45). **(g)**-**(i)** Performance comparison in the middle-age group (45 ≤ *yr <* 75). **(j)**-**(l)** Performance comparison in the old-age group (*yr* ≥ 75).

**Extended Figure S14:**
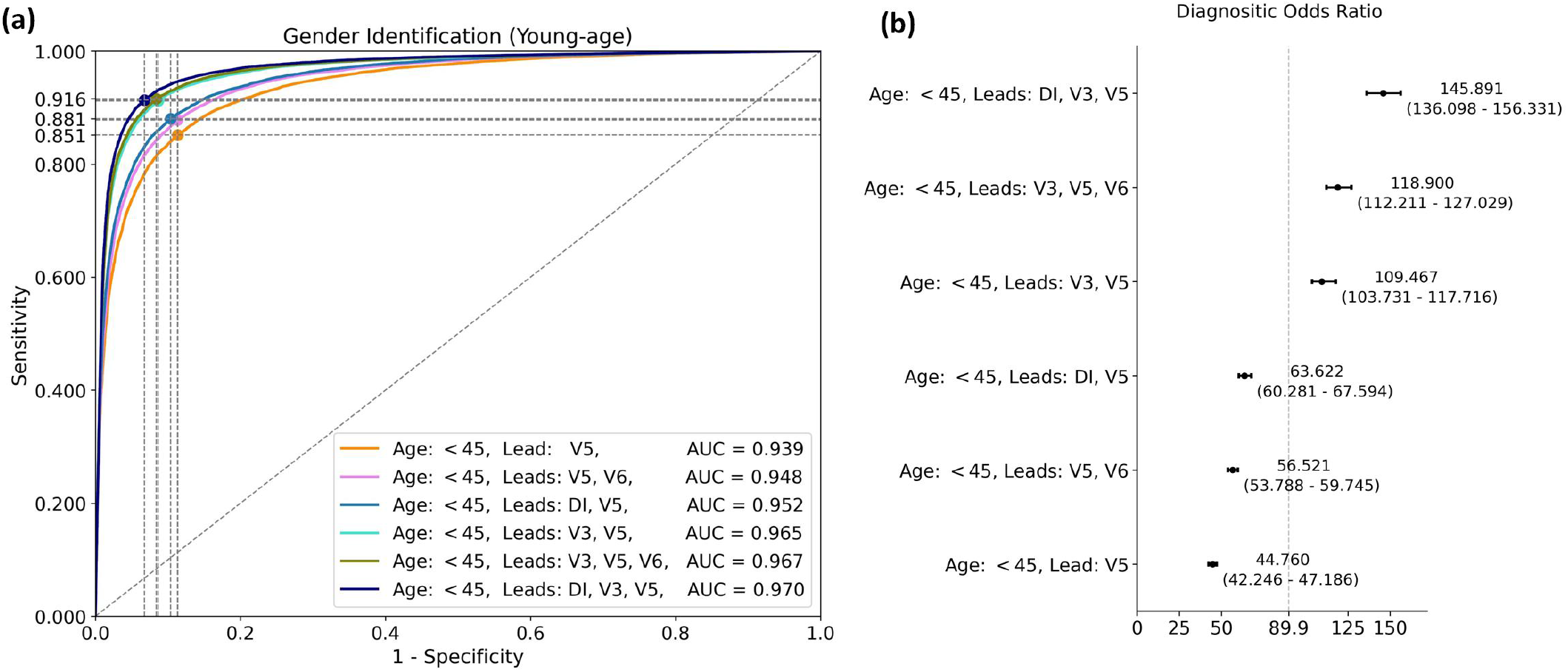
Model performance on gender identification using dominant ECG leads for young-age group (*yr <* 45).**(a)** The ROC and AUC scores for gender identification using different combinations of dominant leads. **(b)** The distribution of DOR values (95% CI) for model performance on gender identification using different combinations of dominant leads.

**Extended Table S5:** Performance comparison of gender identification for young-age group (*yr <* 45) using dominant leads.

| Dominant ECG Leads | Sensitivity<br>(95% CI) | Specificity<br>(95% CI) | AUC Score<br>(95% CI) |
| --- | --- | --- | --- |
| Lead: V5 | 0.851<br>(0.845-0.878) | 0.886<br>(0.860-0.893) | 0.939<br>(0.936-0.940) |
| Leads: V5, V6 | 0.878<br>(0.871-0.884) | 0.887<br>(0.881-0.894) | 0.948<br>(0.946-0.950) |
| Leads: DI, V5 | 0.881<br>(0.876-0.891) | 0.896<br>(0.886-0.901) | 0.952<br>(0.950-0.953) |
| Leads: V3, V5 | 0.912<br>(0.899-0.924) | 0.914<br>(0.902-0.927) | 0.965<br>(0.964-0.967) |
| Leads: V3, V5, V6 | 0.916<br>(0.909-0.920) | 0.916<br>(0.912-0.923) | 0.967<br>(0.965-0.968) |
| Leads: DI, V3, V5 | 0.914<br>(0.910-0.930) | 0.932<br>(0.918-0.936) | 0.970<br>(0.969-0.972) |

**Extended Figure S15:**
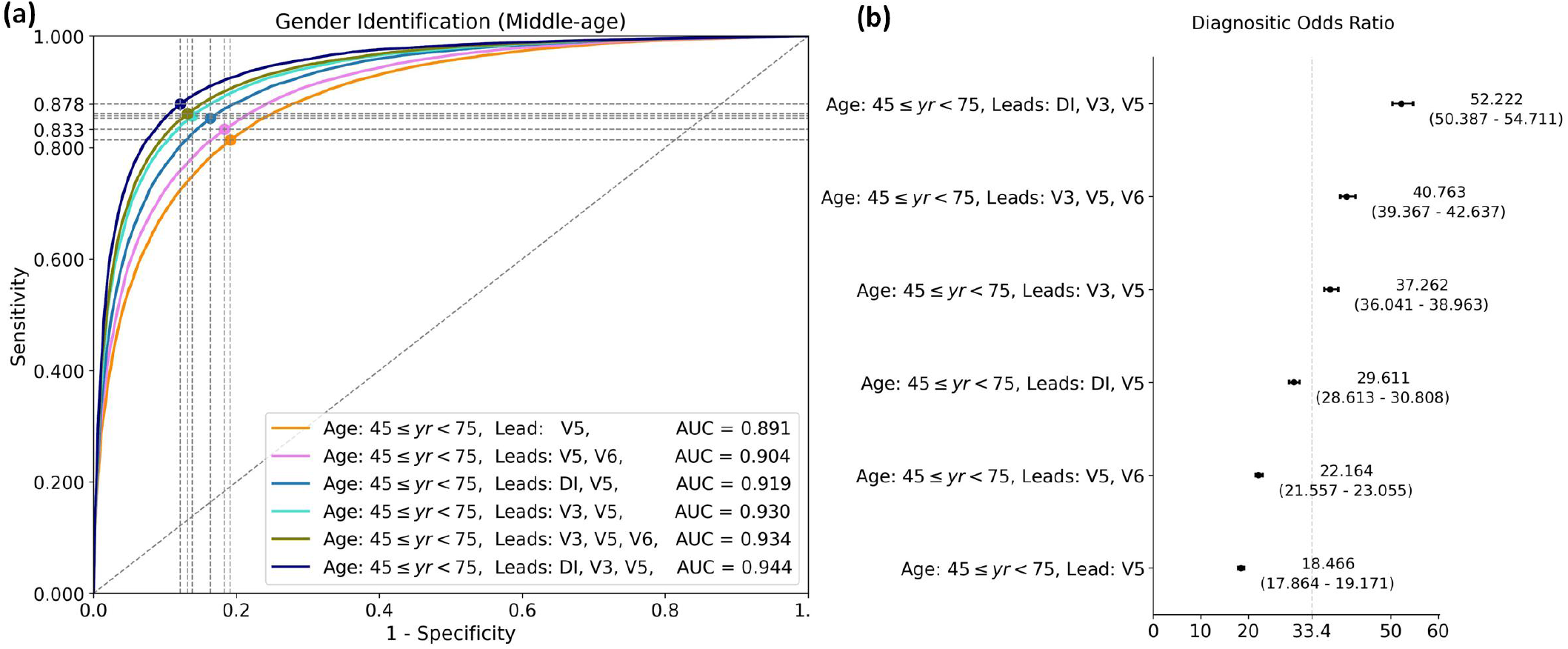
Model performance of gender identification using dominant ECG leads for middle-age group (45 ≤ *yr <* 75). **(a)** The ROC and AUC scores for gender identification using different combinations of dominant ECG leads. **(b)** The distribution of DOR values (95% CI) for model performance on gender identification using different combinations of dominant leads.

**Extended Table S6:** Performance comparison of gender identification for middle-age group (45 ≤ *yr <* 75) using dominant ECG leads.

| Dominant ECG Leads | Sensitivity<br>(95% CI) | Specificity<br>(95% CI) | AUC Score<br>(95% CI) |
| --- | --- | --- | --- |
| Lead: V5 | 0.814<br>(0.807-0.822) | 0.809<br>(0.801-0.815) | 0.891<br>(0.888-0.893) |
| Leads: V5, V6 | 0.833<br>(0.805-0.836) | 0.817<br>(0.815-0.844) | 0.904<br>(0.902-0.906) |
| Leads: DI, V5 | 0.853<br>(0.835-0.860) | 0.837<br>(0.829-0.854) | 0.919<br>(0.917-0.921) |
| Leads: V3, V5 | 0.857<br>(0.844-0.867) | 0.862<br>(0.852-0.875) | 0.930<br>(0.928-0.932) |
| Leads: V3, V5, V6 | 0.861<br>(0.848-0.872) | 0.868<br>(0.858-0.882) | 0.934<br>(0.933-0.936) |
| Leads: DI, V3, V5 | 0.878<br>(0.869-0.886) | 0.879<br>(0.871-0.888) | 0.944<br>(0.942-0.945) |

**Extended Figure S16:**
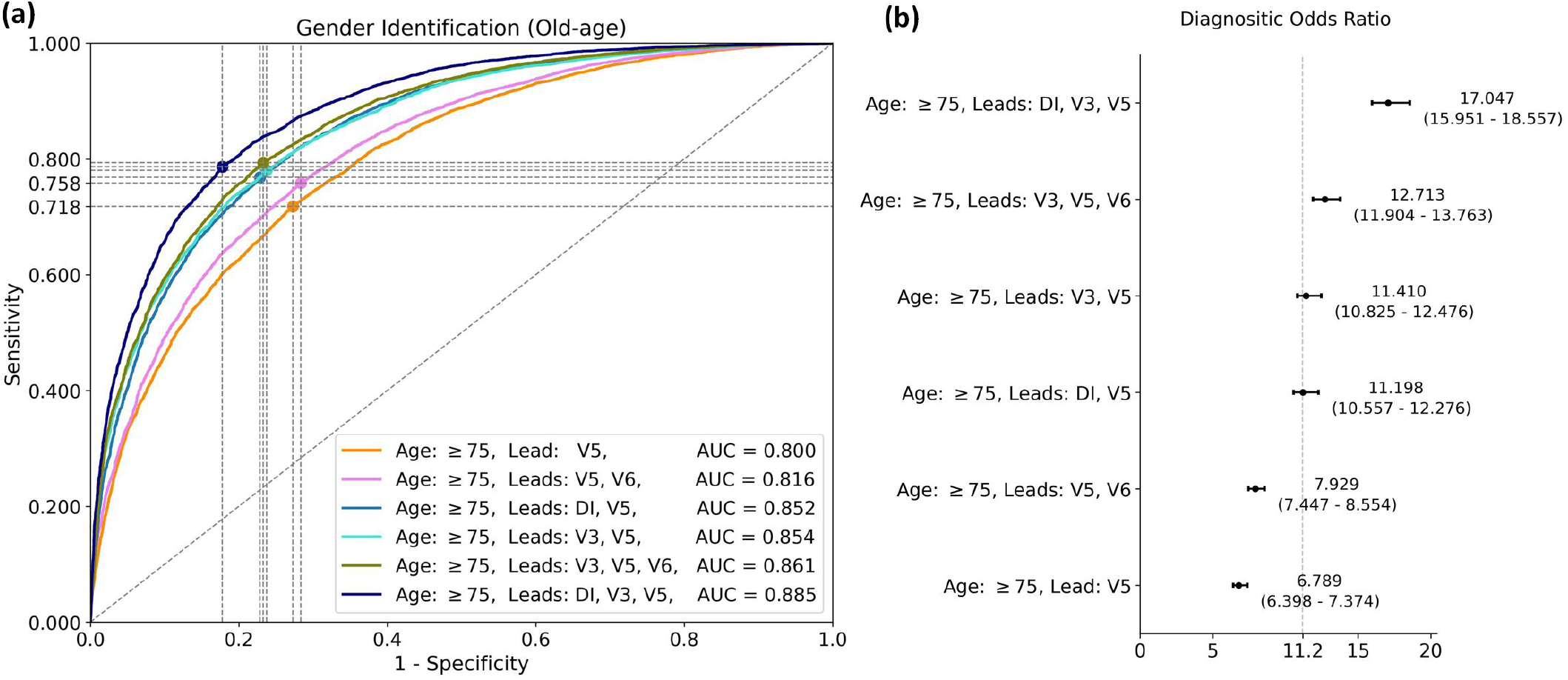
Model performance of gender identification using dominant ECG leads for old-age group (*yr >* 75). **(a)** The ROC and AUC scores for gender identification using different combinations of dominant leads. **(b)** The distribution of DOR values (95% CI) for model performance on gender identification using different combinations of dominant leads.

**Extended Table S7:** Performance comparison of gender identification for old-age group (*yr* ≥ 75) using dominant leads.

| Dominant ECG Leads | Sensitivity<br>(95% CI) | Specificity<br>(95% CI) | AUC Score<br>(95% CI) |
| --- | --- | --- | --- |
| Lead: V5 | 0.718<br>(0.705-0.753) | 0.727<br>(0.694-0.741) | 0.800<br>(0.793-0.806) |
| Leads: V5, V6 | 0.758<br>(0.716-0.777) | 0.716<br>(0.700-0.759) | 0.816<br>(0.809-0.822) |
| Leads: DI, V5 | 0.768<br>(0.756-0.820) | 0.771<br>(0.723-0.784) | 0.852<br>(0.846-0.858) |
| Leads: V3, V5 | 0.780<br>(0.732-0.800) | 0.763<br>(0.745-0.813) | 0.854<br>(0.849-0.860) |
| Leads: V3, V5, V6 | 0.794<br>(0.762-0.803) | 0.767<br>(0.759-0.798) | 0.861<br>(0.856-0.866) |
| Leads: DI, V3, V5 | 0.786<br>(0.781-0.834) | 0.822<br>(0.777-0.828) | 0.885<br>(0.880-0.890) |

**Extended Figure S17:**
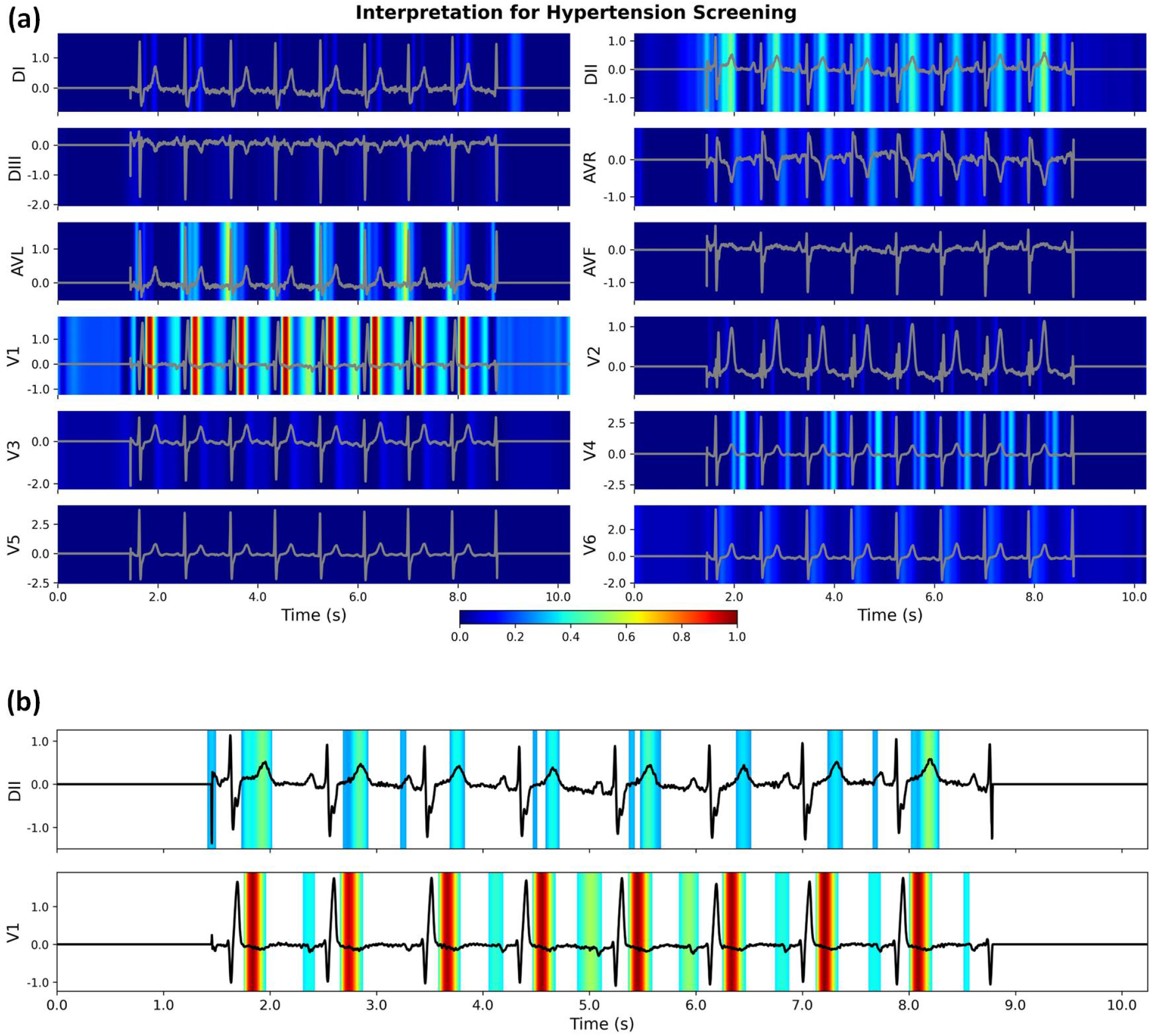
Interpretation for hypertension screening using our developed CResNet model. **(a)** The original calculated heatmaps for 12 ECG leads, with colour bar ranging from blue to red indicating the increasing weights of importance. **(b)** The refined view of the DII and V1 leads with background colour removed. Using the combination of salient features, the CResNet model has a probability of 0.902 to screen hypertension for the subject using the ECG recording.

**Extended Figure S18:**
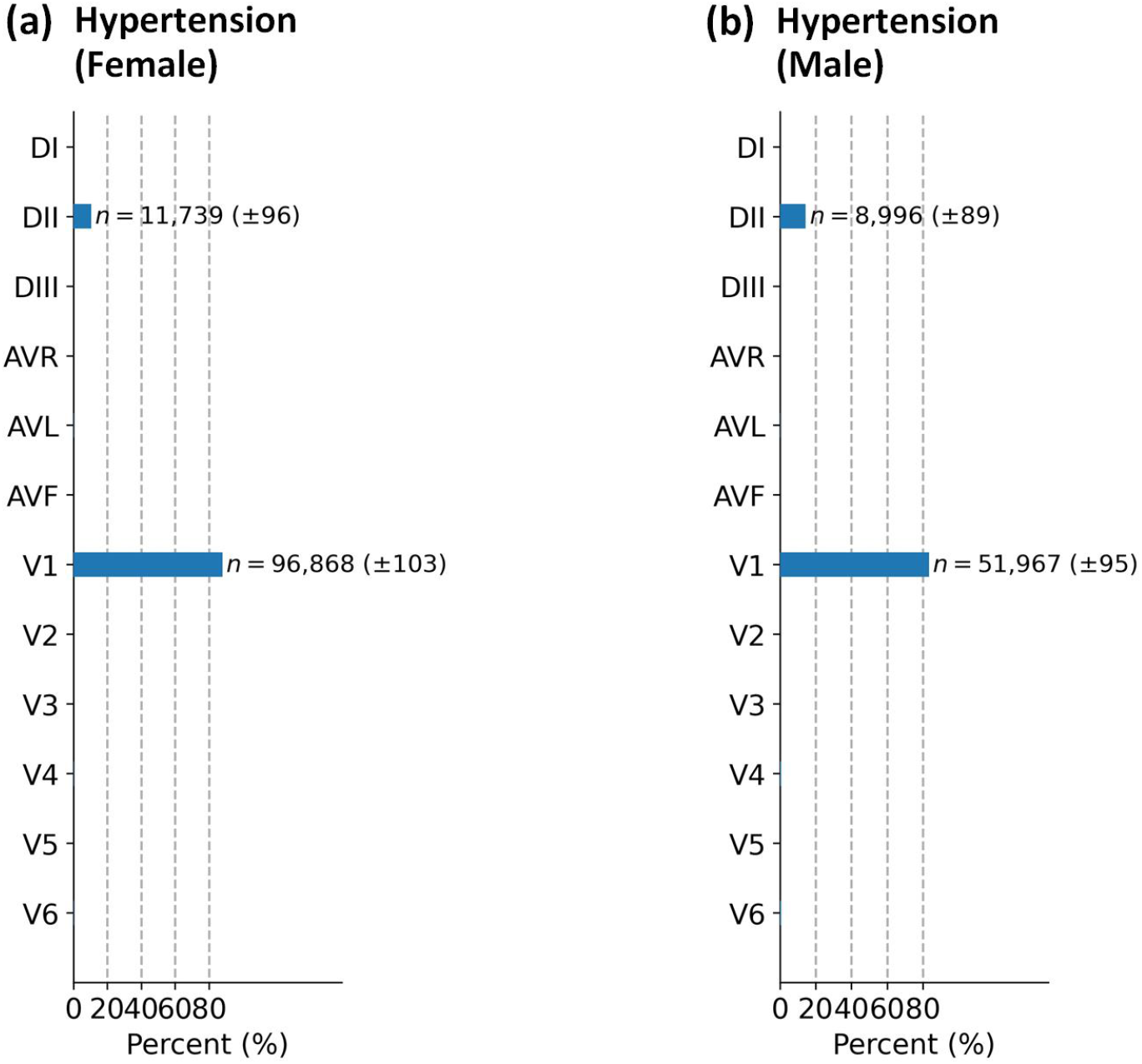
Distributions of dominant ECG leads for hypertension screening using our developed CResNet model. **(a)** Distribution of dominant leads for female subjects. **(b)** Distribution of dominant leads for male subjects. We annotate the number of occurrences when the dominant lead accounts for more than 10% of all the 12 ECG leads. The number of occurrences is presented as mean and standard deviation.

**Extended Figure S19:**
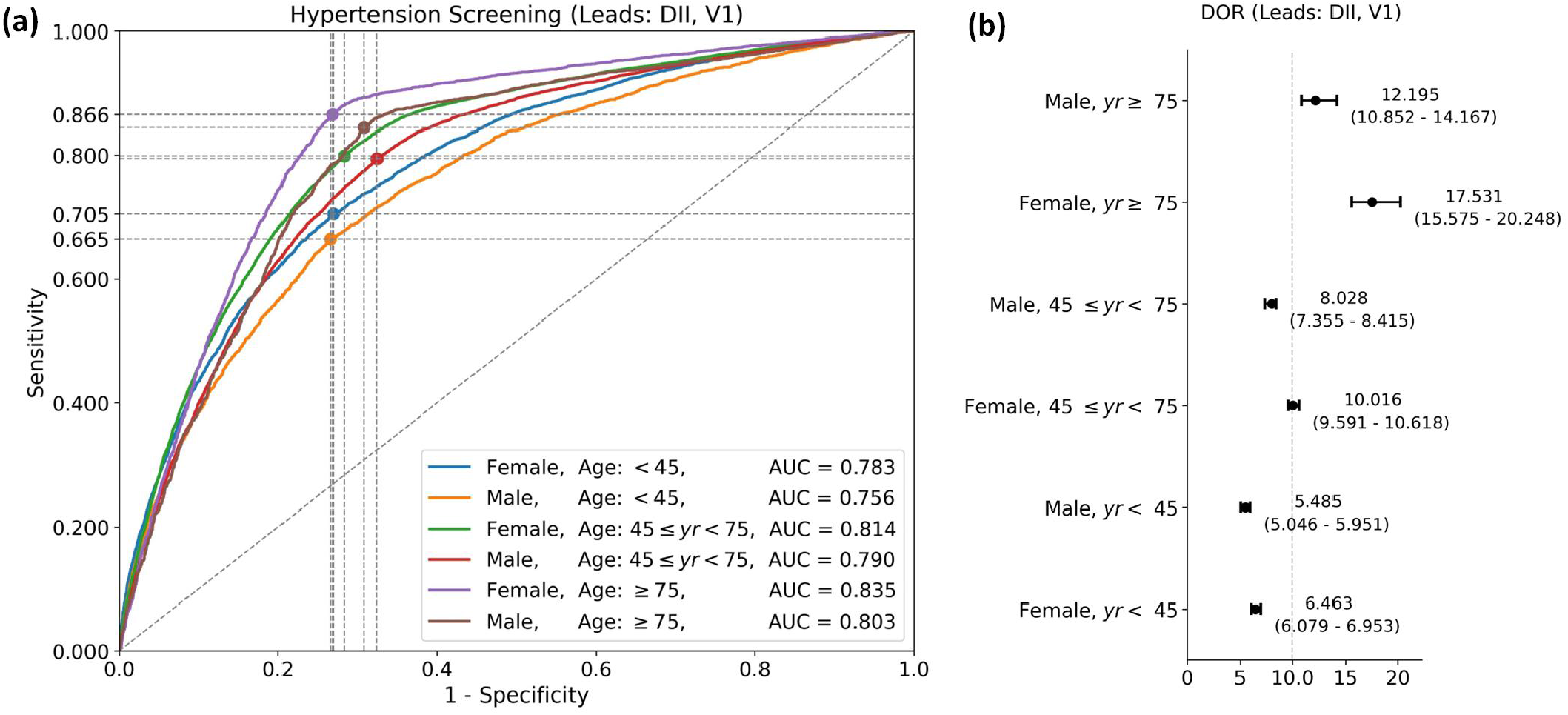
Model performance on hypertension screening in different populations using the dominant DII and V1 leads. **(a)** The ROC and AUC scores for hypertension screening in different populations. **(b)** The distribution of DOR values (95% CI) for model performance on hypertension screening in different populations.

**Extended Table S8:**
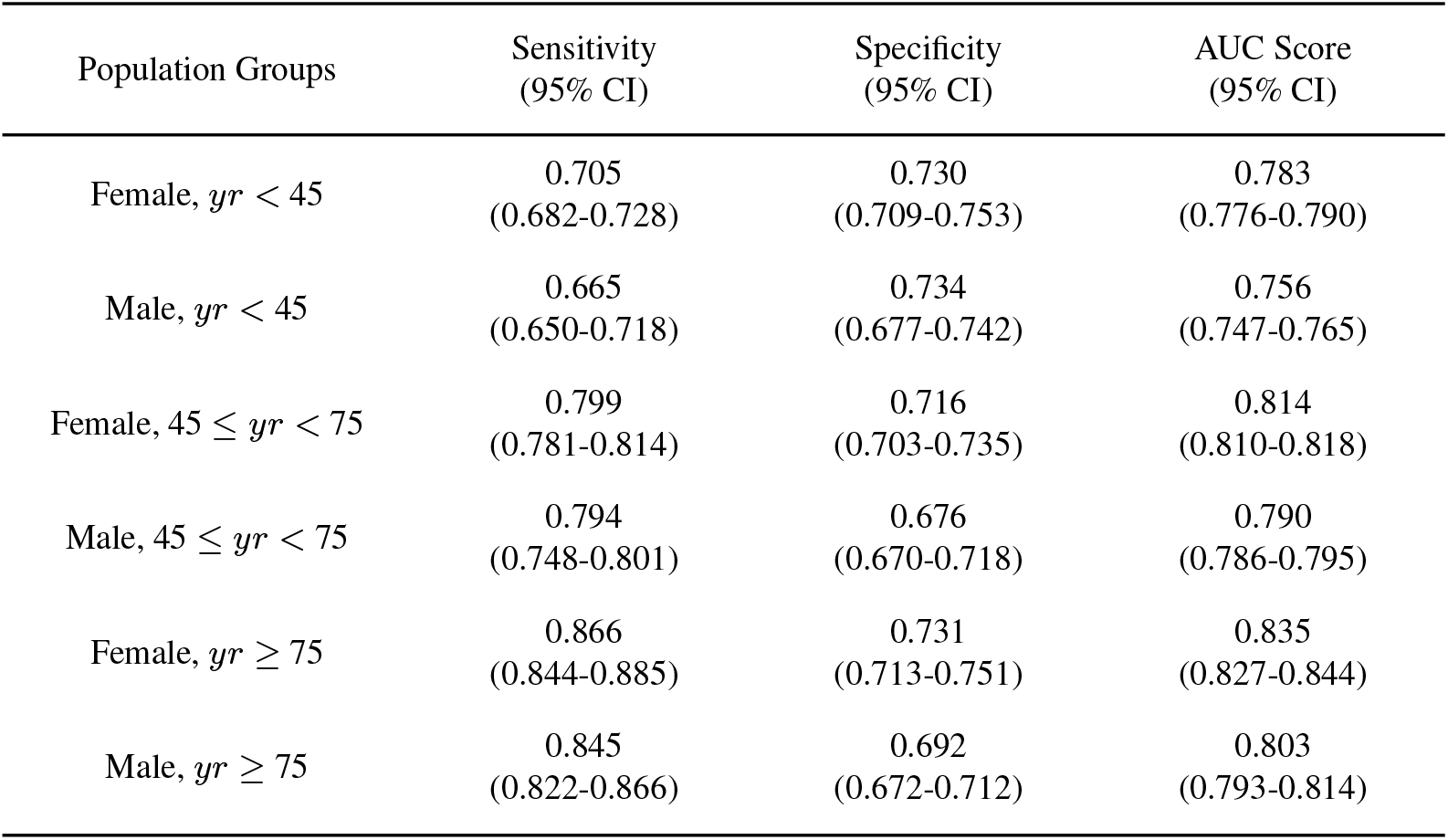
Performance comparison for hypertension screening using dominant DII and V1 ECG leads in terms of age and gender differences.

**Extended Figure S20:**
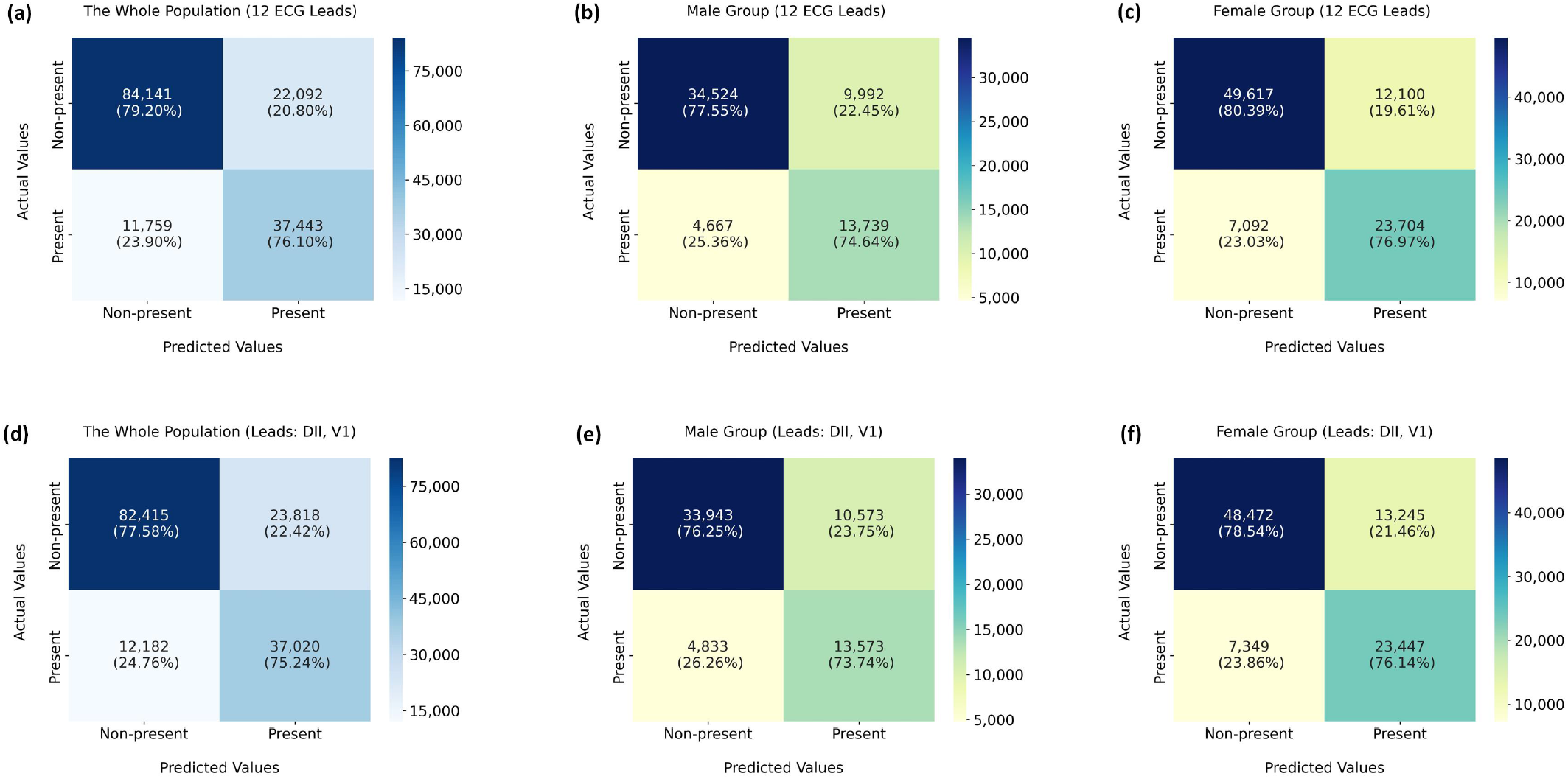
Confusion matrices for hypertension screening using 12 ECG leads, and the dominant DII and V1 leads. **(a)**-**(c)** Performance comparison of hypertension screening using 12-lead ECGs in different populations. **(d)**-**(f)** Performance comparison of hypertension screening using dominant ECG leads in different populations.

**Extended Figure S21:**
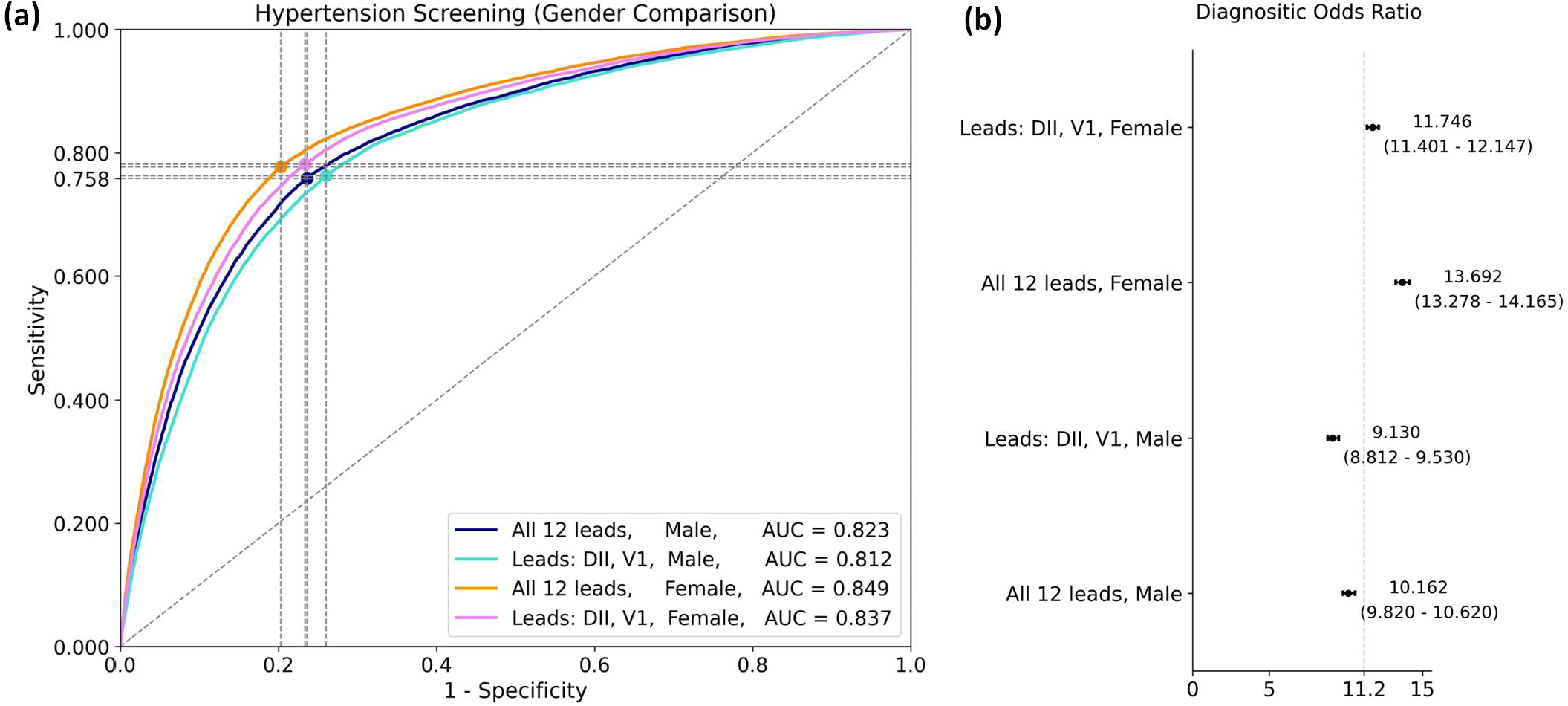
Model performance on hypertension screening using 12 ECG leads and dominant ECG leads in terms of gender differences. **(a)** The ROC and AUC scores for hypertension screening using 12 ECG leads and dominant ECG leads. **(b)** The distribution of DOR values (95% CI) for model performance on hypertension screening using 12 ECG leads and dominant ECG leads.

**Extended Table S9:** Performance comparison on hypertension screening using different ECG leads in terms of gender differences.

| ECG Leads & Gender Differences | Sensitivity<br>(95% CI) | Specificity<br>(95% CI) | AUC Score<br>(95% CI) |
| --- | --- | --- | --- |
| All 12 leads, Male | 0.758<br>(0.745-0.772) | 0.764<br>(0.752-0.778) | 0.823<br>(0.820-0.827) |
| Leads: DII, V1, Male | 0.763<br>(0.744-0.775) | 0.740<br>(0.729-0.760) | 0.812<br>(0.808-0.816) |
| All 12 leads, Female | 0.777<br>(0.769-0.791) | 0.797<br>(0.784-0.805) | 0.849<br>(0.847-0.852) |
| Leads: DII, V1, Female | 0.781<br>(0.765-0.787) | 0.767<br>(0.763-0.783) | 0.837<br>(0.834-0.840) |

**Extended Figure S22:**
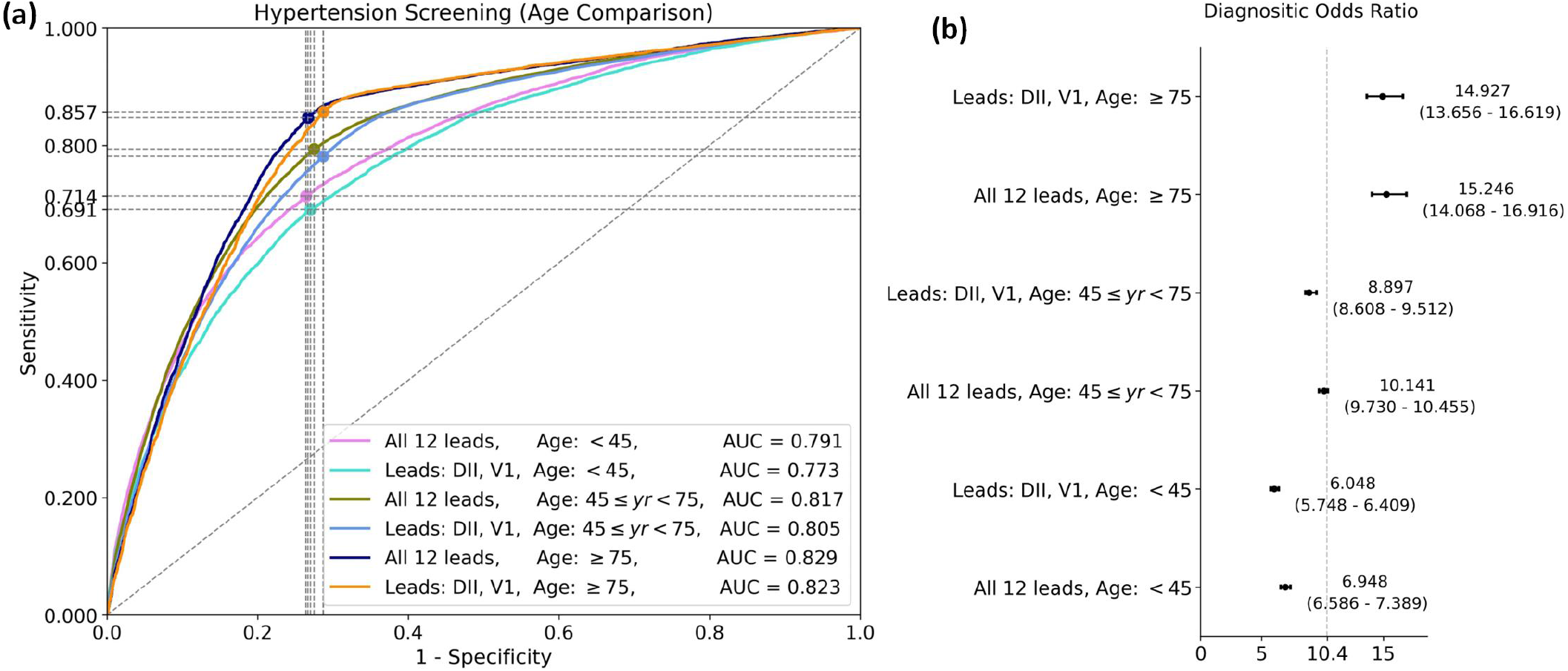
Model performance on hypertension screening using 12 ECG leads and dominant ECG leads in terms of age differences. **(a)** The ROC and AUC scores for hypertension screening using 12 ECG leads and dominant ECG leads. **(b)** The distribution of DOR values (95% CI) for model performance on hypertension screening using 12 ECG leads and dominant ECG leads.

**Extended Table S10:** Performance comparison on hypertension screening using different ECG leads in terms of age differences.

| ECG Leads & Age Differences | Sensitivity<br>(95% CI) | Specificity<br>(95% CI) | AUC Score<br>(95% CI) |
| --- | --- | --- | --- |
| All 12 leads, Age: $< 45$ | 0.714<br>(0.687-0.736) | 0.736<br>(0.717-0.763) | 0.791<br>(0.785-0.796) |
| Leads: DII, V1, Age: $< 45$ | 0.691<br>(0.678-0.711) | 0.730<br>(0.710-0.740) | 0.773<br>(0.767-0.779) |
| All 12 leads, Age: $45 \leq yr < 75$ | 0.794<br>(0.774-0.798) | 0.725<br>(0.721-0.744) | 0.817<br>(0.814-0.820) |
| Leads: DII, V1, Age: $45 \leq yr < 75$ | 0.782<br>(0.770-0.809) | 0.713<br>(0.690-0.724) | 0.805<br>(0.802-0.808) |
| All 12 leads, Age: $\geq 75$ | 0.847<br>(0.831-0.865) | 0.733<br>(0.717-0.749) | 0.829<br>(0.822-0.836) |
| Leads: DII, V1, Age: $\geq 75$ | 0.857<br>(0.844-0.874) | 0.713<br>(0.697-0.726) | 0.823<br>(0.816-0.829) |

**Extended Figure S23:**
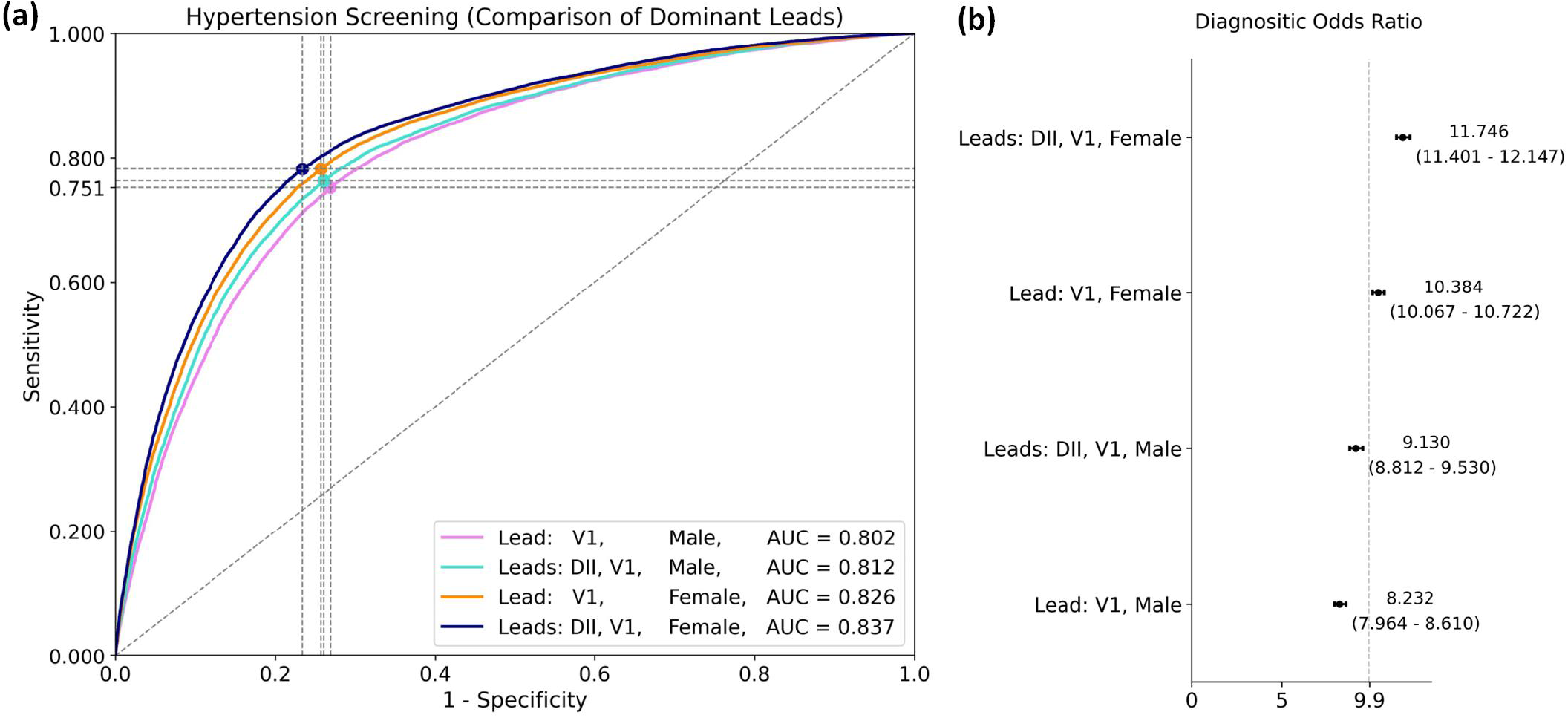
Model performance on hypertension screening using different combinations of dominant ECG leads in terms of gender differences. **(a)** The ROC and AUC scores for hypertension screening using different dominant ECG leads. **(b)** The distribution of DOR values (95% CI) for model performance on hypertension screening using different dominant ECG leads.

**Extended Table S11:** Performance comparison for hypertension screening using different dominant ECG leads in terms of gender differences.

| ECG Leads & Gender Differences | Sensitivity<br>(95% CI) | Specificity<br>(95% CI) | AUC Score<br>(95% CI) |
| --- | --- | --- | --- |
| Lead: V1, Male | 0.751<br>(0.738-0.764) | 0.731<br>(0.719-0.746) | 0.802<br>(0.798-0.806) |
| Leads: DII, V1, Male | 0.763<br>(0.744-0.775) | 0.740<br>(0.729-0.760) | 0.812<br>(0.808-0.816) |
| Lead: V1, Female | 0.782<br>(0.751-0.793) | 0.743<br>(0.733-0.773) | 0.826<br>(0.823-0.829) |
| Leads: DII, V1, Female | 0.781<br>(0.765-0.787) | 0.767<br>(0.763-0.783) | 0.837<br>(0.834-0.840) |

## References

[1] Boris Babic et al. “Beware explanations from AI in health care”. Science 373.6552 (2021), pp. 284–286.

[2] Stephanie L Hyland et al. “Early prediction of circulatory failure in the intensive care unit using machine learning”. Nature Medicine 26.3 (2020), pp. 364–373.

[3] Faiz Ahmad Khan et al. “Chest X-ray analysis with deep learning-based software as a triage test for pulmonary tubercu-losis: a prospective study of diagnostic accuracy for culture-confirmed disease”. The Lancet Digital Health 2.11 (2020), e573–e581.

[4] Jeffrey De Fauw et al. “Clinically applicable deep learning for diagnosis and referral in retinal disease”. Nature Medicine 24.9 (2018), pp. 1342–1350.

[5] Sushravya Raghunath et al. “Prediction of mortality from 12-lead electrocardiogram voltage data using a deep neural network”. Nature Medicine 26.6 (2020), pp. 886–891.

[6] Alvaro E Ulloa Cerna et al. “Deep-learning-assisted analysis of echocardiographic videos improves predictions of allcause mortality”. Nature Biomedical Engineering 5.6 (2021), pp. 546–554.

[7] Andre Esteva et al. “Dermatologist-level classification of skin cancer with deep neural networks”. Nature 542.7639 (2017), pp. 115–118.

[8] Yiqiu Shen et al. “Artificial intelligence system reduces false-positive findings in the interpretation of breast ultrasound exams”. Nature Communications 12.1 (2021), pp. 1–13.

[9] Varun Gulshan et al. “Development and validation of a deep learning algorithm for detection of diabetic retinopathy in retinal fundus photographs”. JAMA 316.22 (2016), pp. 2402–2410.

[10] Eric J Topol. “High-performance medicine: the convergence of human and artificial intelligence”. Nature Medicine 25.1 (2019), pp. 44–56.

[11] Yonatan Elul et al. “Meeting the unmet needs of clinicians from AI systems showcased for cardiology with deep-learning– based ECG analysis”. Proceedings of the National Academy of Sciences 118.24 (2021).

[12] Pranav Rajpurkar et al. “AI in health and medicine”. Nature Medicine (2022), pp. 1–8.

[13] US Food and Drug Administration. “Artificial intelligence/machine learning (AI/ML)-based software as a medical device (SAMD) action plan”. US Food Drug Administration, White Oak, MD, USA, Technical Report 145022 (2021).

[14] Shinjini Kundu. “AI in medicine must be explainable”. Nature Medicine 27.8 (2021), pp. 1328–1328.

[15] Yujin Oh, Sangjoon Park, and Jong Chul Ye. “Deep learning COVID-19 features on CXR using limited training data sets”. IEEE Transactions on Medical Imaging 39.8 (2020), pp. 2688–2700.

[16] Xueyi Zheng et al. “Deep learning radiomics can predict axillary lymph node status in early-stage breast cancer”. Nature Communications 11.1 (2020), pp. 1–9.

[17] Bolei Zhou et al. “Learning deep features for discriminative localization”. In: Proceedings of IEEE Conference on Computer Vision and Pattern Recognition. 2016, pp. 2921–2929.

[18] Marco Tulio Ribeiro, Sameer Singh, and Carlos Guestrin. ““Why should I trust you?” Explaining the predictions of any classifier”. In: Proceedings of the 22nd ACM SIGKDD International Conference on Knowledge Discovery and Data Mining. 2016, pp. 1135–1144.

[19] Scott M Lundberg and Su-In Lee. “A unified approach to interpreting model predictions”. Advances in Neural Information Processing Systems 30 (2017).

[20] Ramprasaath R Selvaraju et al. “Grad-CAM: Visual explanations from deep networks via gradient-based localization”. Proceedings of IEEE International Conference on Computer Vision (2017), pp. 618–626.

[21] Michael W Sjoding et al. “Deep learning to detect acute respiratory distress syndrome on chest radiographs: a retrospective study with external validation”. The Lancet Digital Health 3.6 (2021), e340–e348.

[22] Alfred P Yoon et al. “Development and validation of a deep learning model using convolutional neural networks to identify scaphoid fractures in radiographs”. JAMA Network Open 4.5 (2021), e216096–e216096.

[23] Aurore Lyon et al. “Computational techniques for ECG analysis and interpretation in light of their contribution to medical advances”. Journal of The Royal Society Interface 15.138 (2018), p. 20170821.

[24] Rafael Ortega et al. “Electrocardiographic monitoring in adults”. New England Journal of Medicine 372.8 (2015), e11.

[25] Hongling Zhu et al. “Automatic multilabel electrocardiogram diagnosis of heart rhythm or conduction abnormalities with deep learning: a cohort study”. The Lancet Digital Health 2.7 (2020), e348–e357.

[26] Awni Y Hannun et al. “Cardiologist-level arrhythmia detection and classification in ambulatory electrocardiograms using a deep neural network”. Nature Medicine 25.1 (2019), pp. 65–69.

[27] Zachi I Attia et al. “Screening for cardiac contractile dysfunction using an artificial intelligence–enabled electrocardiogram”. Nature Medicine 25.1 (2019), pp. 70–74.

[28] Michal Cohen-Shelly et al. “Electrocardiogram screening for aortic valve stenosis using artificial intelligence”. European Heart Journal 42.30 (2021), pp. 2885–2896.

[29] Xiaoxi Yao et al. “Artificial intelligence–enabled electrocardiograms for identification of patients with low ejection fraction: a pragmatic, randomized clinical trial”. Nature Medicine 27.5 (2021), pp. 815–819.

[30] Zachi I Attia et al. “An artificial intelligence-enabled ECG algorithm for the identification of patients with atrial fibrillation during sinus rhythm: a retrospective analysis of outcome prediction”. The Lancet 394.10201 (2019), pp. 861–867.

[31] Jeya Vikranth Jeyakumar et al. “How can I explain this to you? an empirical study of deep neural network explanation methods”. Advances in Neural Information Processing Systems 33 (2020), pp. 4211–4222.

[32] James H Thrall et al. “Artificial intelligence and machine learning in radiology: opportunities, challenges, pitfalls, and criteria for success”. Journal of the American College of Radiology 15.3 (2018), pp. 504–508.

[33] Amy Groenewegen et al. “Epidemiology of heart failure”. European Journal of Heart Failure 22.8 (2020), pp. 1342–1356.

[34] Rola El-Serag and Rebecca C Thurston. “Matters of the heart and mind: interpersonal violence and cardiovascular disease in women”. Journal of the American Heart Association 9.4 (2020), e015479.

[35] Michael E Mendelsohn and Richard H Karas. “Molecular and cellular basis of cardiovascular gender differences”. Science 308.5728 (2005), pp. 1583–1587.

[36] Naomi DL Fisher and Gregory Curfman. “Hypertension—a public health challenge of global proportions”. JAMA 320.17 (2018), pp. 1757–1759.

[37] Katherine T Mills, Andrei Stefanescu, and Jiang He. “The global epidemiology of hypertension”. Nature Reviews Nephrology 16.4 (2020), pp. 223–237.

[38] Antônio H Ribeiro et al. “Automatic diagnosis of the 12-lead ECG using a deep neural network”. Nature Communications 11.1 (2020), pp. 1–9.

[39] Peter A Noseworthy et al. “Artificial intelligence-guided screening for atrial fibrillation using electrocardiogram during sinus rhythm: a prospective non-randomised interventional trial”. The Lancet (2022).

[40] Robert H. Peter, J. J. Morris, and Henry D. Mcintosh. “Relationship of fibrillatory waves and P waves in the electrocar-diogram”. Circulation 33 (1966), pp. 599–606.

[41] David Amar et al. “Autonomic changes preceding the onset of postoperative atrial fibrillation”. Journal of the American College of Cardiology 41.6S1 (2003), pp. 101–101.

[42] Borys Surawicz and Timothy Knilans. Chou’s electrocardiography in clinical practice: adult and pediatric. Elsevier Health Sciences, 2008.

[43] Gari D Clifford et al. “AF classification from a short single lead ECG recording: The PhysioNet/computing in cardiology challenge 2017”. 2017 Computing in Cardiology (CinC) (2017), pp. 1–4.

[44] Zachi I Attia et al. “Age and sex estimation using artificial intelligence from standard 12-lead ECGs”. Circulation: Arrhythmia and Electrophysiology 12.9 (2019), e007284.

[45] Rajesh Tota-Maharaj et al. “Coronary artery calcium for the prediction of mortality in young adults < 45 years old and elderly adults > 75 years old”. European Heart Journal 33.23 (2012), pp. 2955–2962.

[46] Rutger R van de Leur et al. “Discovering and visualizing disease-specific electrocardiogram features using deep learning: proof-of-concept in phospholamban gene mutation carriers”. Circulation: Arrhythmia and Electrophysiology 14.2 (2021), e009056.

[47] Praharsh Ivaturi et al. “A comprehensive explanation framework for biomedical time series classification”. IEEE Journal of Biomedical and Health Informatics 25.7 (2021), pp. 2398–2408.

[48] Steven A Hicks et al. “Explaining deep neural networks for knowledge discovery in electrocardiogram analysis”. Scientific Reports 11.1 (2021), pp. 1–11.

[49] Roy M John et al. “Ventricular arrhythmias and sudden cardiac death”. The Lancet 380.9852 (2012), pp. 1520–1529.

[50] John Hampton. The ECG in practice. Churchill Livingstone, 2003.

[51] ABM Abdullah. ECG in medical practice. Jaypee Brothers Medical Publishers, 2014.

[52] ILAN Goldenberg, Arthur J Moss, and Wojciech Zareba. “QT interval: how to measure it and what is “normal”“. Journal of Cardiovascular Electrophysiology 17.3 (2006), pp. 333–336.

[53] So-Ryoung Lee et al. “Blood pressure variability and incidence of new-onset atrial fibrillation: a nationwide population-based study”. Hypertension 75.2 (2020), pp. 309–315.

[54] Furrukh Sana et al. “Wearable devices for ambulatory cardiac monitoring: JACC state-of-the-art review”. Journal of the American College of Cardiology 75.13 (2020), pp. 1582–1592.

[55] Matthew A Reyna et al. “Will two do? Varying dimensions in electrocardiography: the PhysioNet/Computing in Cardiology Challenge 2021”. 2021 Computing in Cardiology (CinC) 48 (2021), pp. 1–4.

## References

[56] Antonio Luiz P Ribeiro et al. “Tele-electrocardiography and bigdata: the CODE (Clinical Outcomes in Digital Electrocar-diography) study”. Journal of Electrocardiology 57 (2019), S75–S78.

[57] Emilly M Lima et al. “Deep neural network estimated electrocardiographic-age as a mortality predictor”. Nature Communications 12 (2021), p. 5117.

[58] Maria Beatriz Alkmim et al. “Improving patient access to specialized health care: the Telehealth Network of Minas Gerais, Brazil”. Bulletin of the World Health Organization 90 (2012), pp. 373–378.

[59] Paul Kligfield et al. “Recommendations for the standardization and interpretation of the electrocardiogram: part I: The electrocardiogram and its technology: a scientific statement from the American Heart Association Electrocardiography and Arrhythmias Committee, Council on Clinical Cardiology; the American College of Cardiology Foundation; and the Heart Rhythm Society: endorsed by the International Society for Computerized Electrocardiology”. Journal of the American College of Cardiology 49.10 (2007), pp. 1109–1127.

[60] Adriano Veloso, Wagner Meira, and Mohammed J Zaki. “Lazy associative classification”. In: Sixth International Conference on Data Mining (ICDM’06). IEEE. 2006, pp. 645–654.

[61] PW Macfarlane et al. “Methodology of ECG interpretation in the Glasgow program”. Methods of Information in Medicine 29.04 (1990), pp. 354–361.

[62] Peter W Macfarlane and Shahid Latif. “Automated serial ECG comparison based on the Minnesota code”. Journal of Electrocardiology 29 (1996), pp. 29–34.

[63] Aditya Chattopadhay et al. “Grad-CAM++: Generalized gradient-based visual explanations for deep convolutional networks”. In: 2018 IEEE Winter Conference on Application of Computer Vision (WACV). IEEE. 2018, pp. 839–847.

[64] Vinod Nair and Geoffrey E. Hinton. “Rectified Linear Units Improve Restricted Boltzmann Machines”. In: Proceedings of the 27th International Conference on International Conference on Machine Learning. ICML’10. Haifa, Israel: Omnipress, 2010, pp. 807–814.

[65] Karen Simonyan and Andrew Zisserman. “Very Deep Convolutional Networks for Large-Scale Image Recognition”. International Conference on Learning Representations abs/1409.1556 (2015).

[66] Diederik P. Kingma and Jimmy Ba. “Adam: A Method for Stochastic Optimization”. 3rd International Conference on Learning Representations, ICLR 2015, San Diego, CA, USA, May 7-9, 2015, Conference Track Proceedings (2015).

[67] Somayeh Sadeghi et al. “Diabetes mellitus risk prediction in the presence of class imbalance using flexible machine learning methods”. BMC Medical Informatics and Decision Making 22.1 (2022), pp. 1–12.

[68] Afina S Glas et al. “The diagnostic odds ratio: a single indicator of test performance”. Journal of Clinical Epidemiology 56.11 (2003), pp. 1129–1135.

[69] Quinn McNemar. “Note on the sampling error of the difference between correlated proportions or percentages”. Psychometrika 12.2 (1947), pp. 153–157.

[70] Maya Varma et al. “Automated abnormality detection in lower extremity radiographs using deep learning”. Nature Machine Intelligence 1.12 (2019), pp. 578–583.

[71] Erping Long et al. “Discrimination of the behavioural dynamics of visually impaired infants via deep learning”. Nature Biomedical Engineering 3.11 (2019), pp. 860–869.

[72] Jacob Cohen. “A coefficient of agreement for nominal scales”. Educational and Psychological Measurement 20.1 (1960), pp. 37–46.

[73] Shreyasi Datta et al. “Identifying normal, AF and other abnormal ECG rhythms using a cascaded binary classifier”. 2017 Computing in Cardiology (CinC) (2017), pp. 1–4.

